# Genetics of transient amnesia highlights a vascular role in memory

**DOI:** 10.1101/2024.08.18.24312185

**Authors:** Manuel A. Rivas, FinnGen, Mark J. Daly

## Abstract

Where memory is stored and how it is processed is poorly understood. Traditionally, genetics of low prevalence traits have been used to get a better understanding of naturally occuring conditions across species. Here, we conduct a genome-wide association of transient global amnesia (TGA), a specific type of heritable amnesia characterized by a sudden, temporary episode of memory loss, across 4,303 patients and over 1 million controls from three population biobanks in Finland, United States, and the United Kingdom, to better understand the genetic basis of memory. We find nine loci associated with TGA (p < 5×10^-8^) including a missense variant in *CRIP1* with high posterior probability (> .9) of being the causally relevant variant using fine-mapping, replicated across 3 cohorts. Furthermore, we find enrichment for a specific cell type, pericyte cells, which are a key part of the neurovascular unit, which includes astrocytes and neurons, and control interactions between neurons and the cerebral vasculature, implicating a direct vascular role in memory formation and retrieval. We anticipate these genetic studies at a larger scale to bring new biological understanding and therapeutic insights into the brain and memory.

## Introduction

Amnesia is a condition that affects memory, involving a loss of memory that may be partial or complete, temporary or permanent. It’s directly related to memory because it specifically impacts the ability to recall information that was previously known or the ability to form new memories. Amnesia primarily impacts explicit (declarative) memory, which involves conscious recall of facts and events. Implicit (nondeclarative) memory, such as skills like riding a bike or swimming, typically remains unaffected because it is thought to be stored in different brain areas^1^. There are several types of amnesia, and they can be caused by various factors including brain injury, disease affecting the brain, certain medications, or psychological trauma. The effects on memory can range from difficulty forming new memories (anterograde amnesia) to trouble recalling past experiences (retrograde amnesia).

Transient global amnesia (TGA) is a specific type of amnesia characterized by a sudden, temporary episode of memory loss, during which the individual cannot form new memories and may have difficulty recalling past events. These episodes are usually brief, typically lasting for several hours, after which the person’s memory gradually returns, and there is no lasting damage to memory^2^. The cause of TGA is not entirely understood, but it’s thought to be related to temporary disruption of blood flow to certain parts of the brain that are involved in memory formation and retrieval. Recent MRI data suggest that a transient perturbation of hippocampal function is the functional correlate of TGA because focal diffusion lesions can be detected in the CA1 field of the hippocampal cornu ammonis^3^. Nonetheless, these data are inconclusive. Notably, a diagnosis of TGA is not given when the amnesia may be related to stroke, epilepsy/seizure or neurodegenerative disease^4^.

Age and migraine are established risk factors to trigger TGA^3^. Historically, genetics has been a powerful approach to gain insights into the causal biology of poorly understood conditions^5^. Here, we use genetic analysis of transient global amnesia from global population biobanks to better understand memory. First, FinnGen has directly genotyped and aggregated phenotype outcomes in over 470,000 individuals from Finland, an isolated population with recent bottlenecks that offers an opportunity for studying genetic variants in complex phenotypes^6^. Second, UK Biobank has genotyped and aggregated medical records and phenotypes from over 500,000 individuals^7,8^ living in the UK. Third, AllofUS has sequenced and aggregated phenotype outcomes in over 240,000 individuals from the United States^9^.

## Results

### Exploration of phenotypes in Finland correlated with transient global amnesia in time

Public summary data from FinRegistry, a research project that utilizes the registry system in Finland to combine health data with a wide range of information from nearly the whole population of Finland, indicates that transient global amnesia is associated with transient ischemic attack (Ji = 1.62). In FinnGen we find consistent signals where transient cerebral ischemic attack is associated with a 3.3-fold increased risk of transient global amnesia (-log10(*P*) = 80.8). Nonetheless, only 453 out of the 3020 matched cases that exhibit transient global amnesia have had an episode of transient cerebral ischemic attack.

### Genome-wide meta-analysis identifies 9 loci associated with transient global amnesia

We undertook a meta-analysis of three transient global amnesia genome-wide association studies (GWAS) from FinnGen (number of cases = 3315, number of controls = 471,699), UK Biobank (number of cases = 692, number of controls = 413,669), and AllofUS (number of cases = 296 cases, number of controls = 124,781). We identified 9 new susceptibility loci meeting genome-wide significance as there has been no prior GWAS study conducted of TGA (see **Figure 1A**, see **Table 1**, *P* < 5 x 10^-8^).

**Figure 1.**
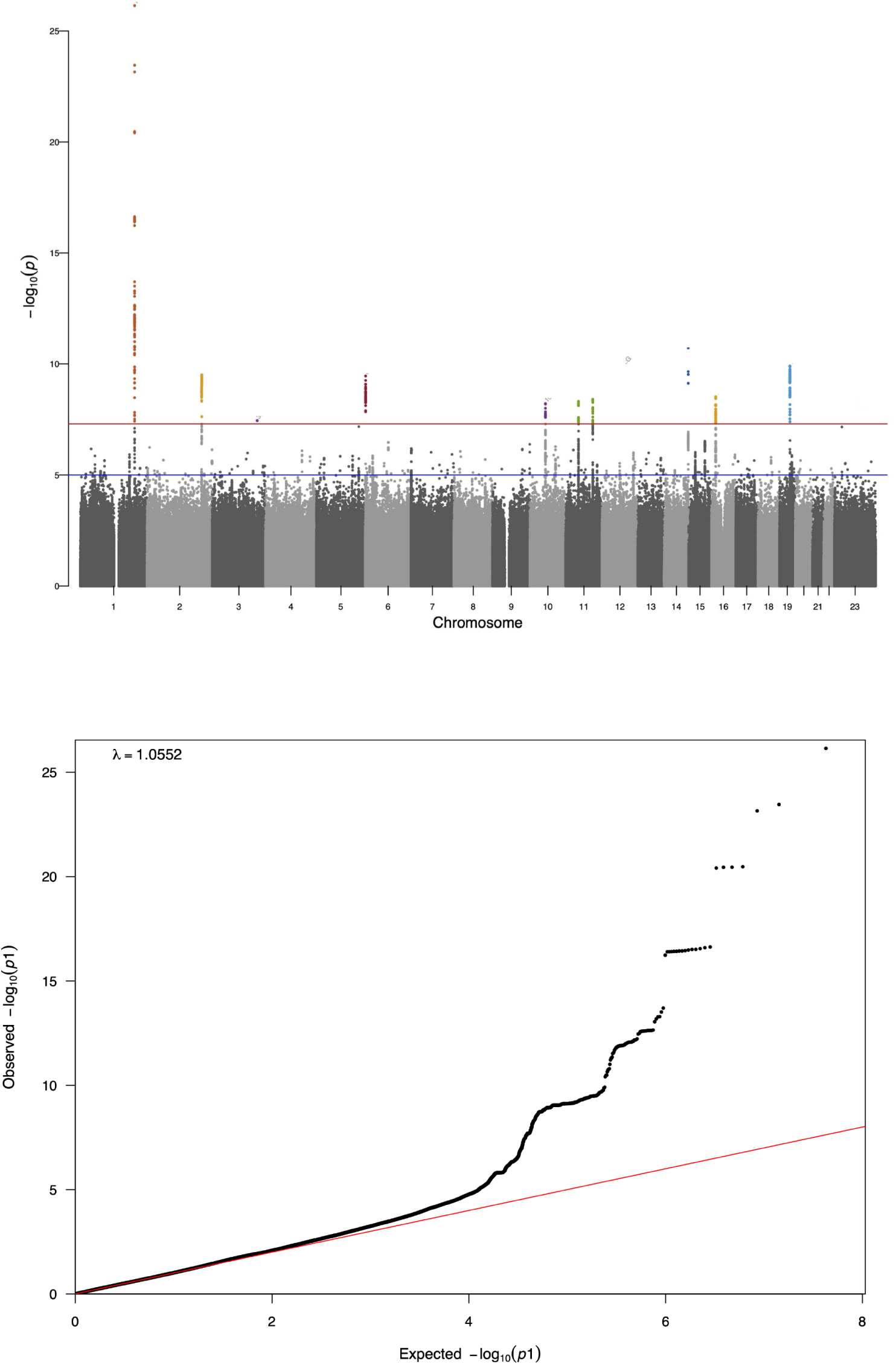
a) Manhattan plot of genome-wide association meta-analysis for transient global amnesia. Summary statistics from GWAS analysis of 3,315 transient global amnesia cases and 471,669 controls in FinnGen were combined with summary statistics from GWAS analysis of 692 TGA cases and 413,669 controls in UK Biobank, and 296 TGA cases and 124,781 controls in AllofUS. (x-axis) genomic position of genetic variant, (y-axis) -log10(P-value) of tested genetic variant. **b) Q-Q plot of p-values from genetic association meta-analysis for transient global amnesia.** (x-axis) -log10(expected P-value) and (y-axis) -log10(observed P-value).

**Table 1.** FinnGen and UK Biobank meta-analysis associated variants with index SNP. Genetic variants with *P* value < 5×10^-8^ are shown. Variant ID corresponds to chromosome-position-ref-alt for hg38 build, rsids column correspond to the rsID of the index variant, nearest genes is the gene proximal to index SNP (when most associated SNPs are localized intrinsically or in a small intergenic region), beta corresponds to estimated log(odds ratio), META_pval correspond to the meta analysis p-value when combining genetic effects estimated in FinnGen and UK Biobank, FINNGEN_af_alt corresponds to the allele frequency of the alternate allele in FinnGen and UKBB_af_alt corresponds to the allele frequency of the alternate allele in UK Biobank. For two loci, associated variants spanned long regions of LD with equivalent evidence for ∼10 genes, and for one (CRIP1) the gene was nominated based on the lead variant being a missense variant. All variants displayed had a *P* value > .05 for heterogeneity of genetic effects, i.e. no significant evidence of heterogeneity observed.

| Variant ID | rsids | nearest_genes | OR | META_pval | Cell type expression |
| --- | --- | --- | --- | --- | --- |
| 1-201926536-T-C | rs72744832 | *LMOD1* | 1.41 | 7.19E-27 | Pericyte cell |
| 14-105488368-C-T | rs55633823 | *CRIP1* | 1.19 | 1.93E-11 | Vascular endothelial cells in cerebellum |
| 19-38694171-G-A | rs7251903 | *ACTN4* | 0.87 | 1.24E-10 | Vascular endothelial cells in cerebellum |
| 2-203331895-C-T | rs116426890 | many | 1.22 | 4.90E-10 |  |
| 6-1366655-G-T | rs192238573 | *FOXF2* | 0.79 | 5.52E-10 | Mural cell brain |
| 16-15814272-T-G | rs12919510 | *MYH11* | 1.14 | 3.00E-09 | Pericyte cell |
| 11-100639014-A-T | rs10894996 | *ARHGAP42* | 0.87 | 4.06E-09 |  |
| 11-47637583-G-A | rs7118178 | many | 1.16 | 4.87E-09 |  |
| 10-58170232-G-A | rs1769016 | *IPMK* | 0.86 | 1.45E-08 |  |

A quantile-quantile plot of the primary meta-statistic using single-SNP z-scores combined across all sample sets showed a marked excess of significant associations (see **Figure 1B**).

### Genetic correlation of transient global amnesia

Genetic correlation estimates are measures that describe the degree of shared genetic influences between two traits or disorders. These estimates can provide insights into whether and how strongly genetic factors contribute to the covariance between traits. Several factors can influence these estimates including pleiotropy, evolutionary history including genetic drift and selective pressures, and ascertainment bias (for example certain individuals may be excluded from the definition of a case thereby giving the appearance that the genetics of a trait has the opposite effect on the trait that was excluded). Here, we estimate the correlation of genetic effects between transient global amnesia and FinnGen phenotypes using LD score regression^10^. LD score regression uses genome-wide data to estimate the correlation of genetic effect parameters thereby not restricting to genome-wide significant findings. We find the genetics of nontraumatic intracranial haemorrhage and migraine to be positively correlated with the genetics of transient global amnesia (rg = 0.29, *P* = .0016; rg = 0.16, *P* = 0.0035, respectively, see **Figure 2**). In addition, we find the genetics of specific development disorders of scholastic skills to be inversely correlated with the genetics of transient global amnesia (rg = -0.48, *P* = .0006). Specific developmental disorders of scholastic skills refers to disorders in which the normal patterns of skill acquisition are disturbed from the early stages of development. It is not simply a consequence of mental retardation, and it is not due to any form of acquired brain trauma or disease^11^. Similarly, the genetics of senile degeneration of the brain and myositis ossificans were inversely correlated with the genetics of TGA (rg = -0.42, *P* = 0.005; rg = -0.33, *P* = .0045, respectively, see **Figure 2**).

**Figure 2.**
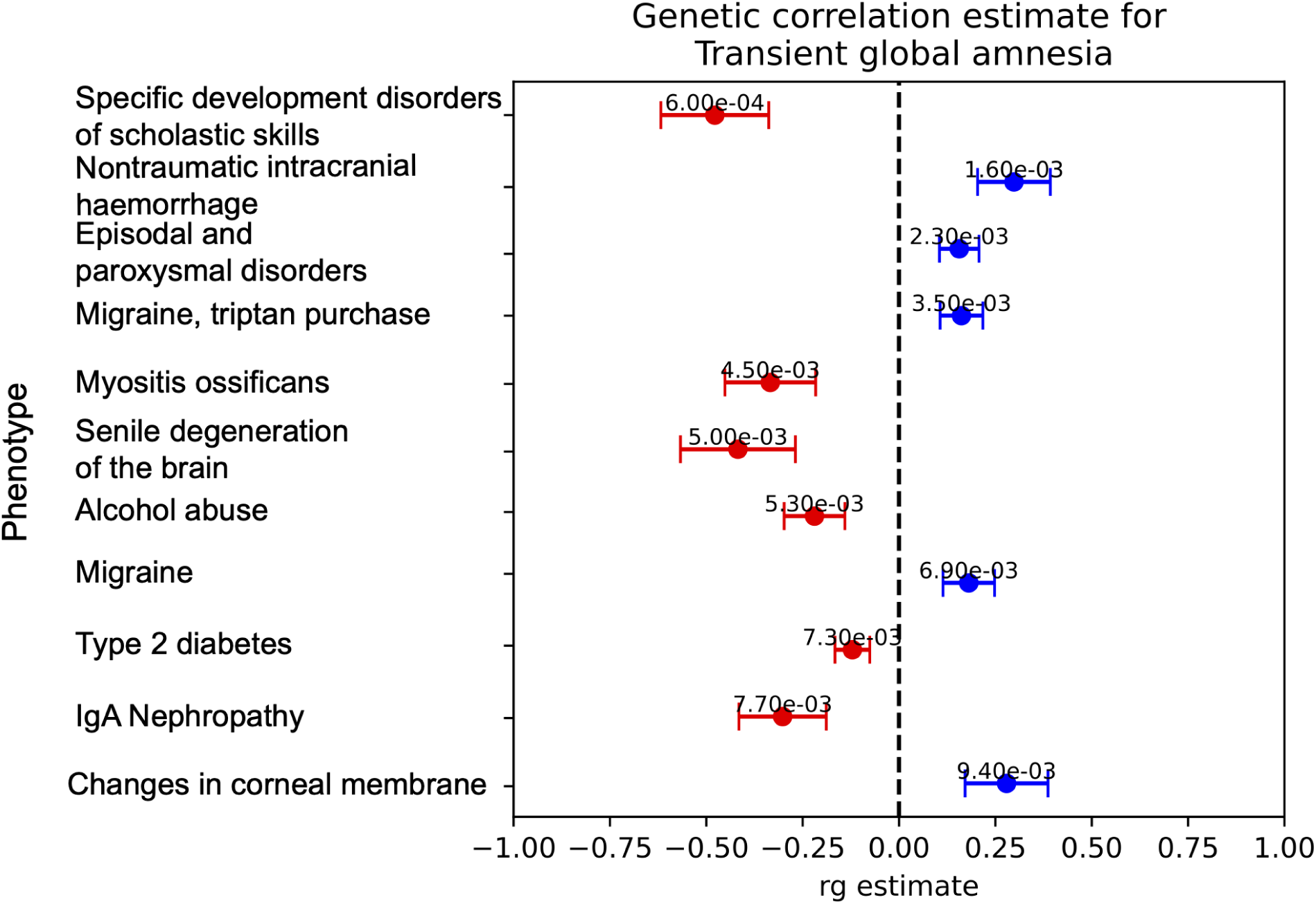
Estimates of the correlation of genetic effects between TGA and other phenotypes across FinnGen and UK Biobank. (x-axis) rg estimate between TGA and (y-axis) phenotype. rg +/- SE is shown, LD score regression *P* value is shown in text. Phenotypes with *P <* .01 are shown.

### Phenome-wide association of 9 TGA lead variants

Given the overall overlap of genetic effects with other conditions we asked whether the top 8 Transient global amnesia associated variants had patterns of pleiotropic effects (assessed as *P* < 5×10^-5^) or whether the effects were specific to TGA. Of the 9 loci, 5 had evidence of being associated with another condition including coronary revascularization and myocardial infarction (n = 2), hypertension (n = 2), and pulse rate and pulse pressure (n=2). Although we did find that the genetic variant proximal to *FOXF2* associated with TGA also associates to stroke, the effects are in the opposite direction (see **Supplementary Table 1**). To our surprise, multiple of the genetic variants that are significantly associated with transient global amnesia are specific, i.e. *P* > 5×10^-5^ across all conditions assessed across both UK Biobank and FinnGen. In particular, we find that the associated variant, rs72744832 (OR = 1.41), an intronic variant in *LMOD1* does not associate with other diseases, but is associated with lean mass phenotypes in UK Biobank. *LMOD1* has high observed expression in tibial, coronary, and aorta artery (median TPM > 800), however quantitative trait analysis does not reveal any specific overlap that may indicate the genetic variant is involved in regulating expression of *LMOD1*^12^. In addition to the cardiovascular phenotypic overlaps being the most prominent, of the 7 genes noted in table 1 as the most likely based on proximity, 4 of these (LMOD1, CRIP1, ACTN4, MYH11) have extremely strong arterial expression in GTEx, while two others (ARHGAP42 and IPMK) are more broadly expressed including substantial arterial expression.

To explore more specifically these potential genes, we used the Enrichr tool^13^ program to probe two recently published compendia of cellular profiles: CellMarker^13,14^ and Tabula Sapiens. The top associations are shown in **Supplementary Table 2 -** of note, the most significant enrichment is with Mural cell brain expression in mouse (p = 9.2e-7, OR = 244) and among human cells in Tabula Sapiens, the three equally enriched cell types (p<.001) include vascular pericyte cells (the subset of mural cells relevant to small vessels/capillaries) (p = 6.7e-4, OR = 67)^13^.

### Protein-altering variant, p.Ala58Val, in *CRIP1* is associated with memory loss

Given established enrichment (along with improved interpretability) of protein-altering variants associated with disease^15,16^ we asked whether any of the significantly associated variants pinpointed protein-altering variants that associate with TGA. Here, we found that a missense variant, p.Ala58Val, in *CRIP1* associates with transient global amnesia (meta-analysis *P* = 2.1×10^-11^, see **Figure 3**) - top index SNP in the region. The genetic variant has consistent effects in UK Biobank (odds ratio = 1.28, *P* = 3.1×10^-5^) and FinnGen (odds ratio = 1.17, *P* = 6.9×10^-8^) highlighting the robustness of the association. *CRIP1* is found to be highly expressed in coronary, tibial, and aorta artery tissue^12^. CRIP1 has a role in zinc absorption and functions as an intracellular zinc transport protein^17,18^. This variant has been previously noted as the most likely causal variant (0.75 < PIP < 1) underlying genome wide associations to increased height and heel bone mineral density and decreased calcium levels in UK Biobank quantitative trait analyses^19^.

**Figure 3.**
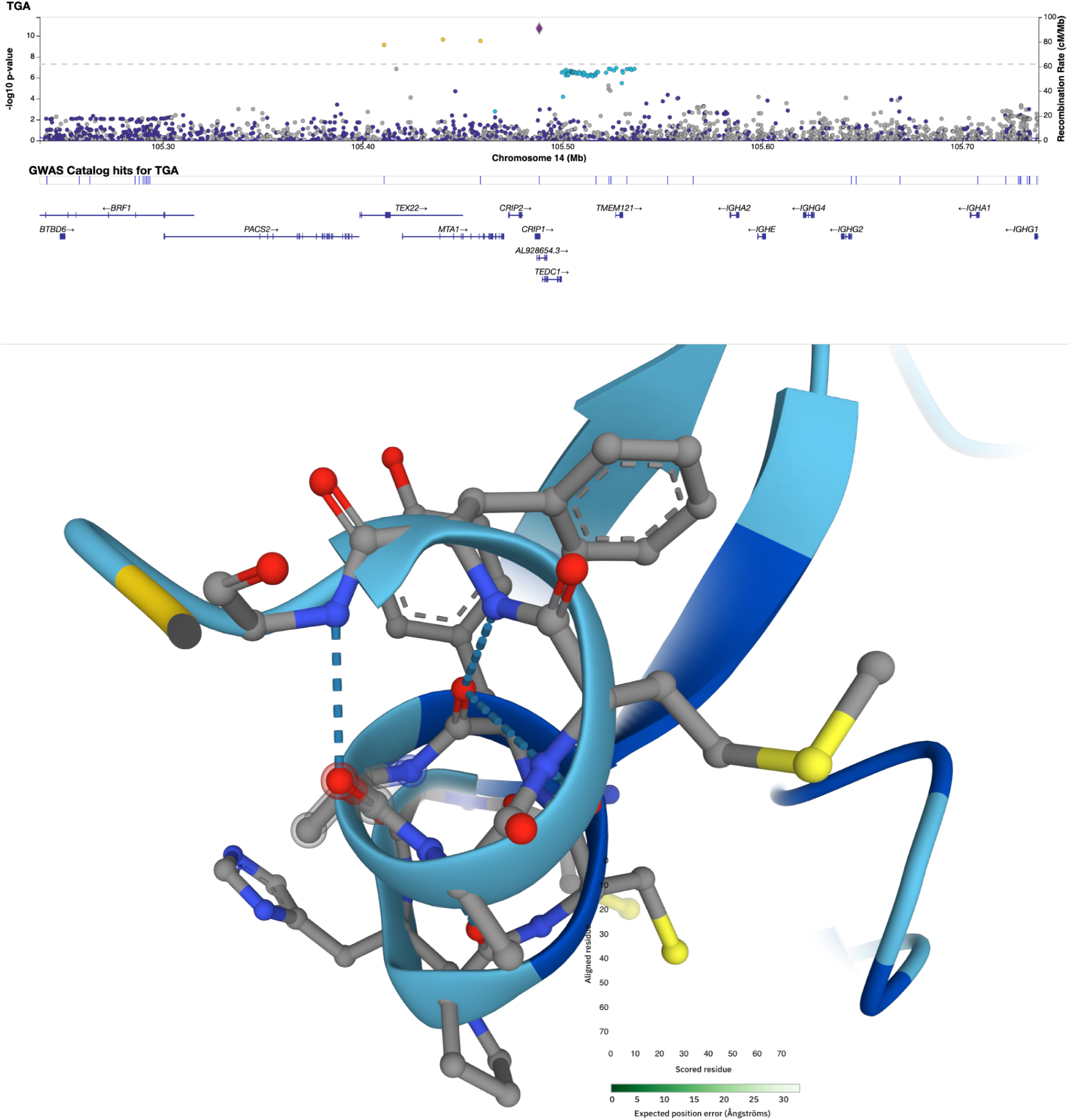
*CRIP1* locus associated with transient global amnesia. On the x-axis is the location and gene diagram of the *CRIP1* locus. The scatter plot shows the associated variants, y-axis corresponds to the -log10(meta analysis *P* value). Highlighted is the missense variant, p.Ala58Val, in *CRIP1*. Location of CRIP1 p.Ala58Val mutation in protein structure UniProt P50238.

## Discussion

In this study we present the first genetic dissection of transient global amnesia, an unusual form of transient memory loss attributable to factors beyond those observed in Alzheimer’s disease or secondary to stroke or seizure. Using genome wide association across three biobanks, totalling 4303 cases and more than 1 million controls, we identify nine genome wide significant associations.

Notably, the variants identified in this study overlap with previously documented associated variants to several cardiovascular phenotypes - with multiple hits to coronary revascularization, hypertension and pulse pressure among the associations with prior evidence of association. Along with a majority of implicated genes showing extremely strong expression in artery, this lends strong support to the hypothesis that TGA may have a largely vascular origin, rather than a neuronal one. The same susceptibility origin has previously been noted for migraine, one of the main established risk factors for TGA, though the associations described here are not broadly here seen in migraine.

One of the nine associations points strongly to a missense variant in CRIP1 that has previously been implicated as the likely causal variant in increased height and bone mineral density along with lower serum calcium levels. We anticipate that these results may open new opportunities towards understanding, in particular, the vascular components of memory and memory loss, providing genes and variants for deeper molecular and phenotypic exploration.

## Data Availability

Summary statistic data are available from the AllofUS workbench, UK Biobank RAP, and FinnGen browser.

## Author information

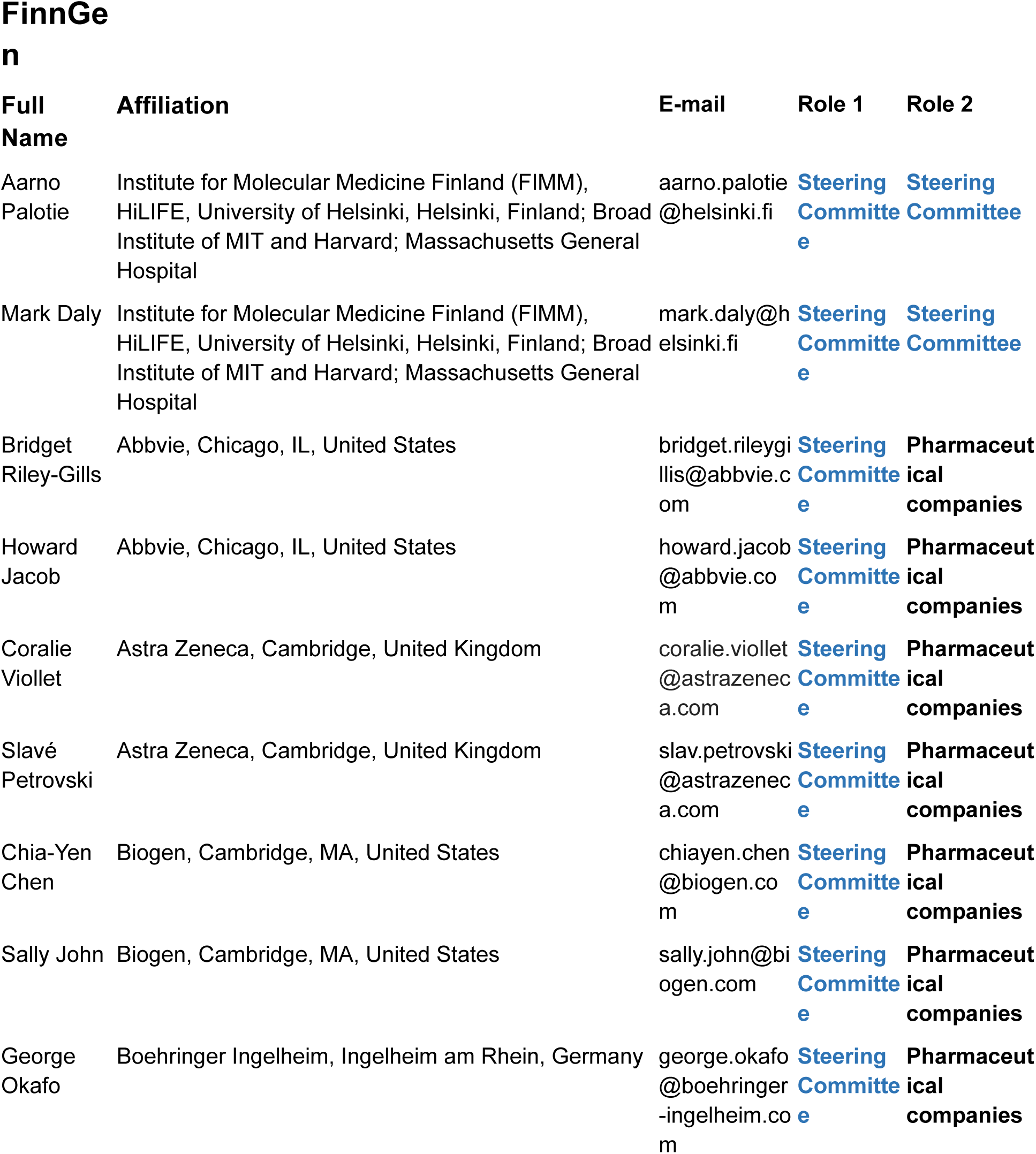
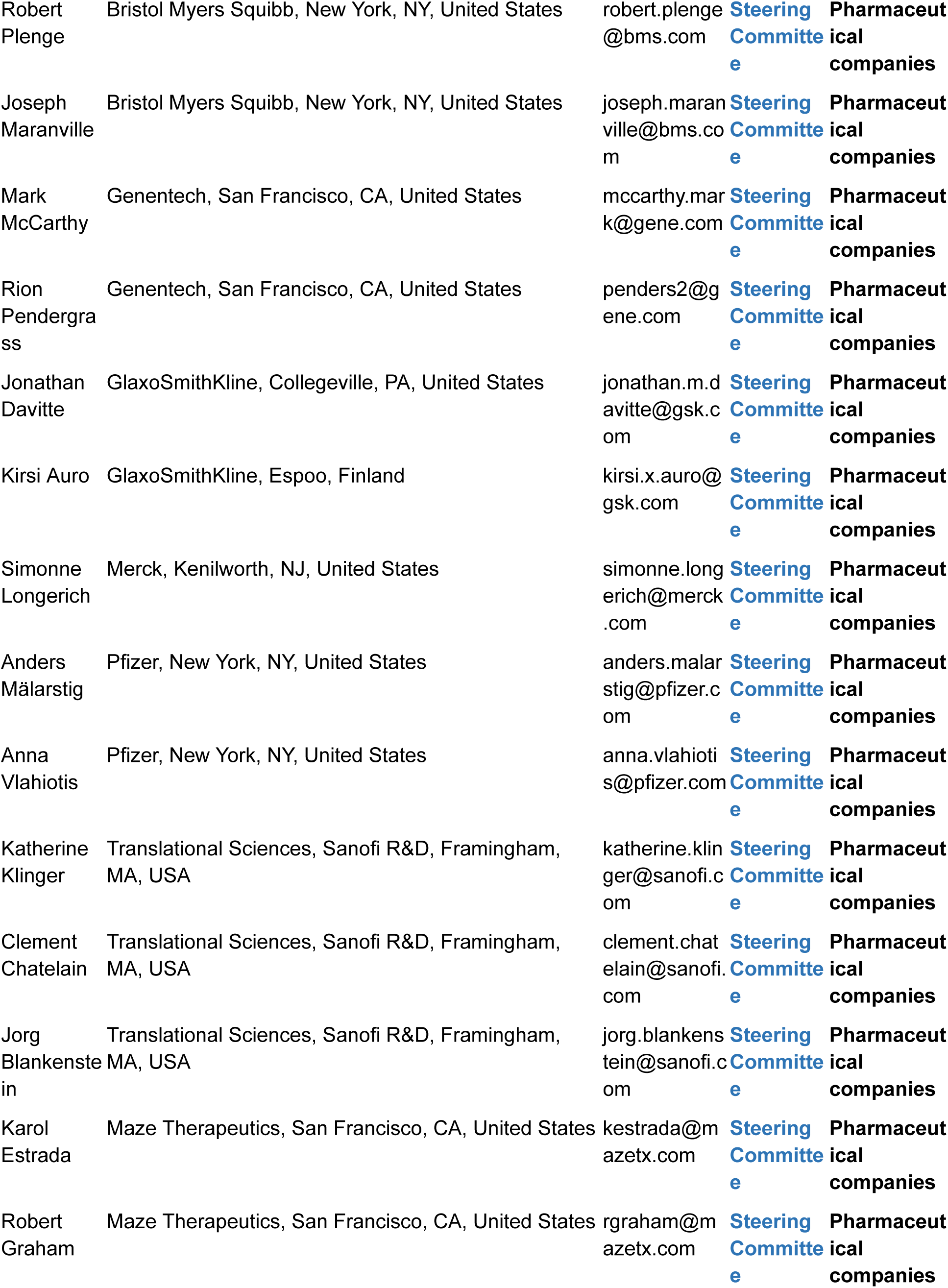
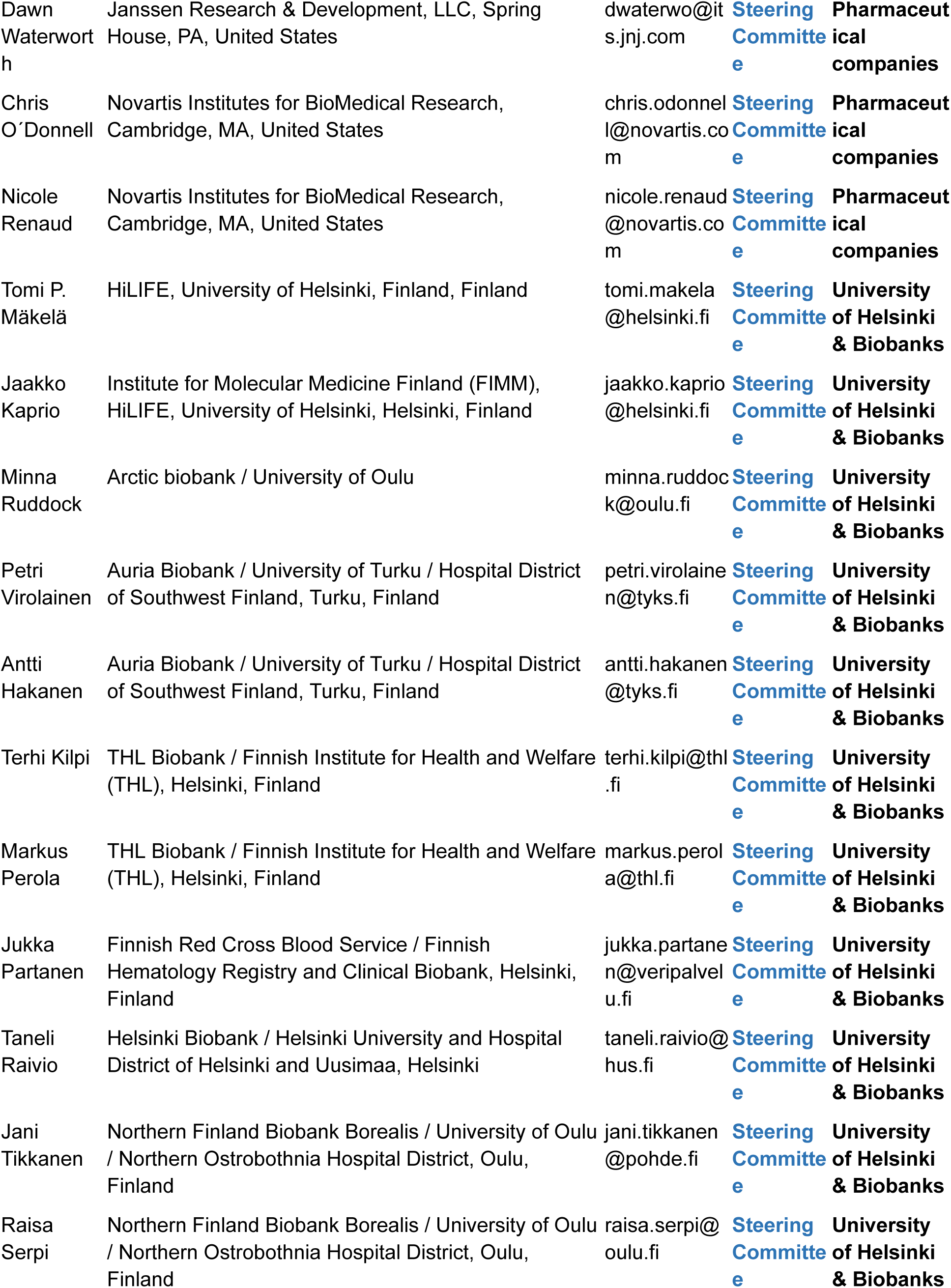
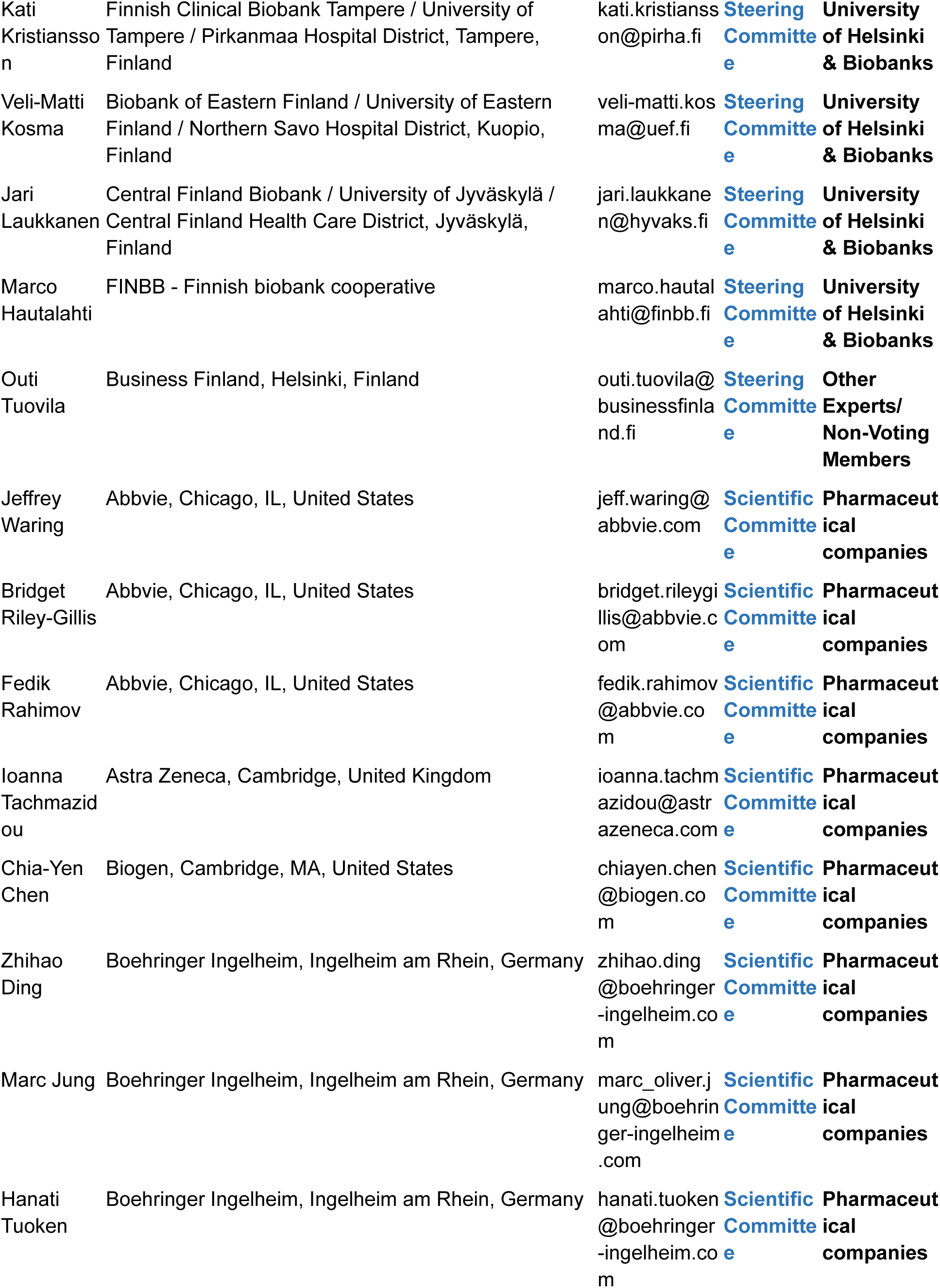
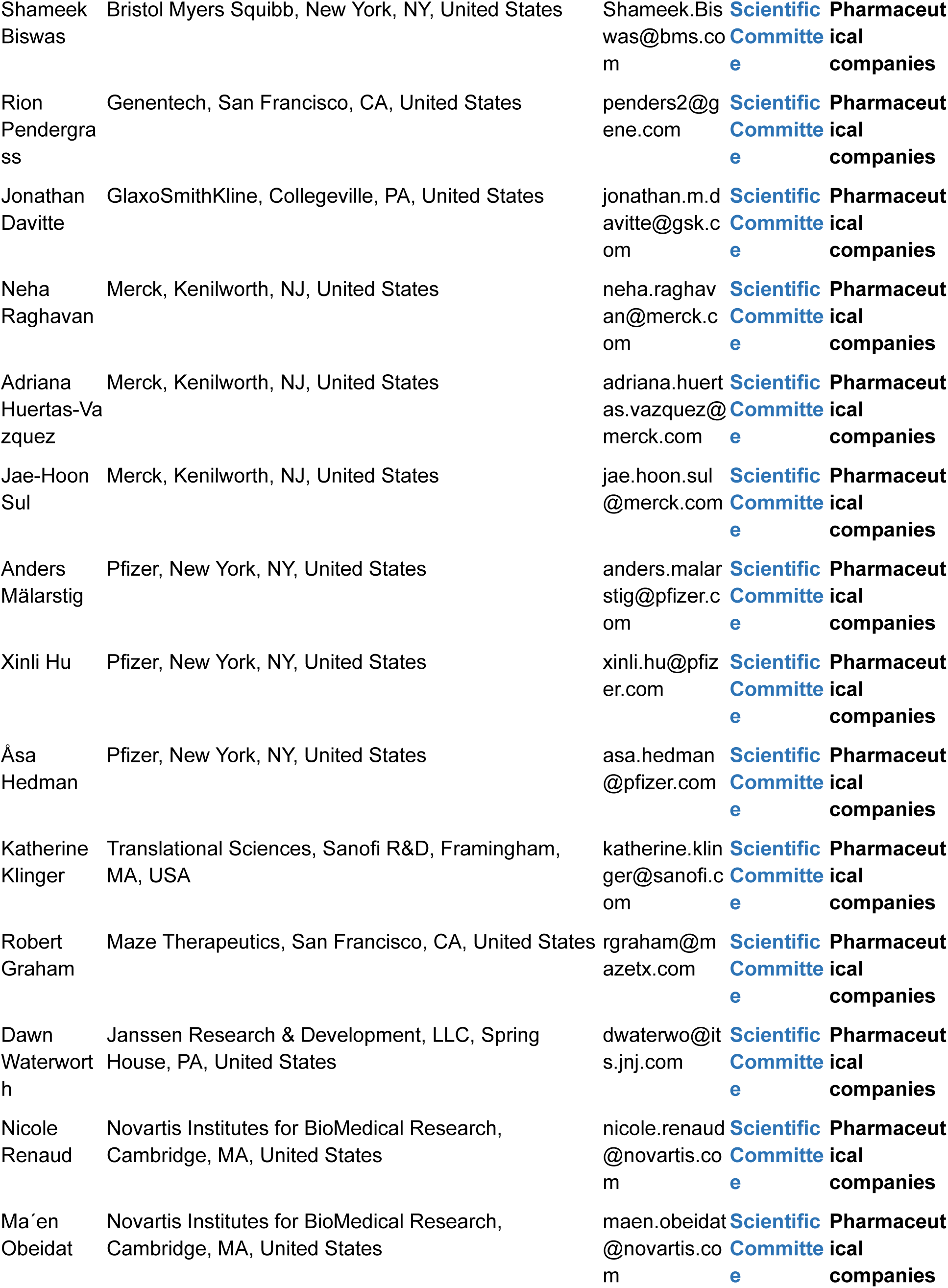
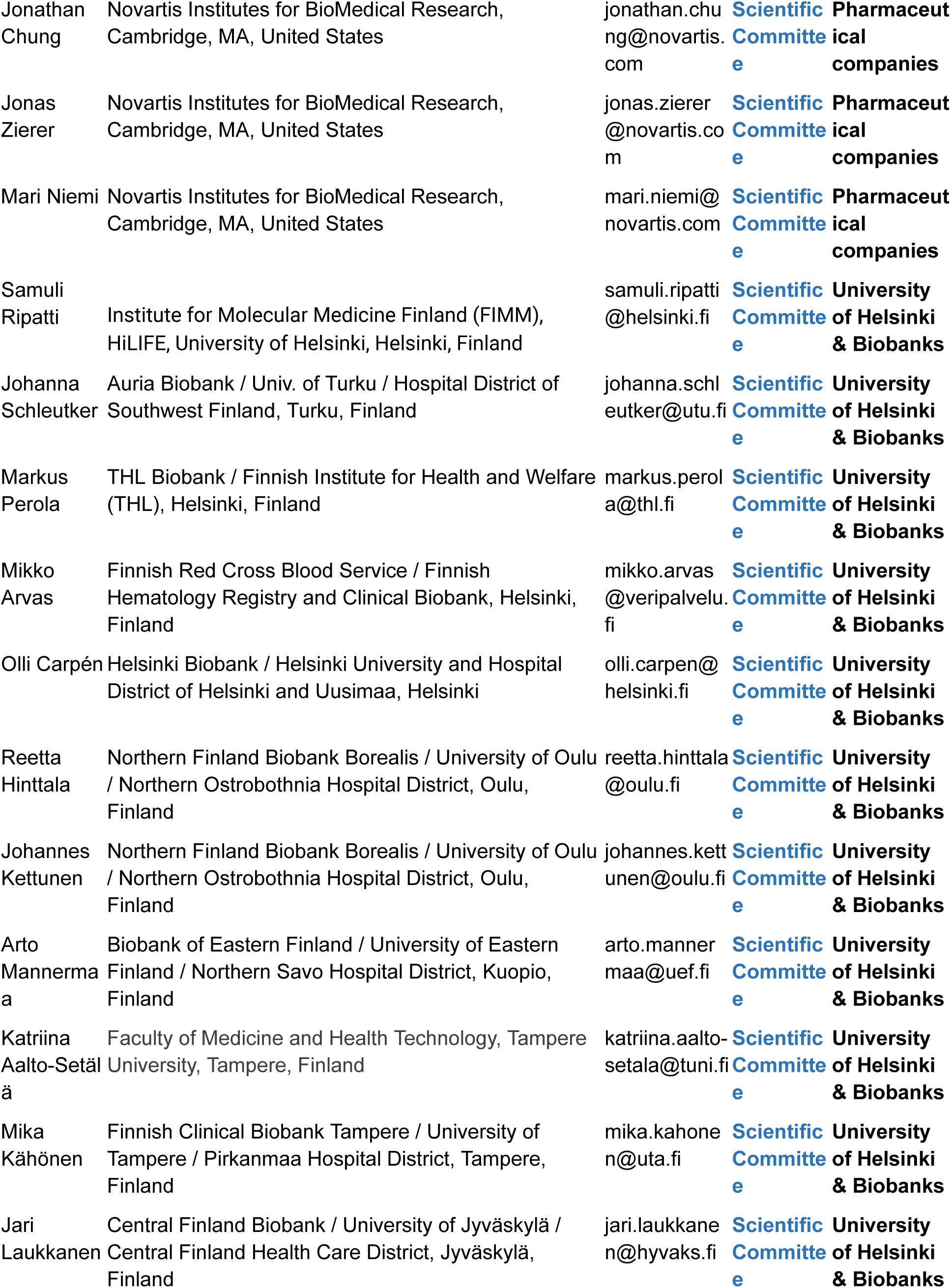
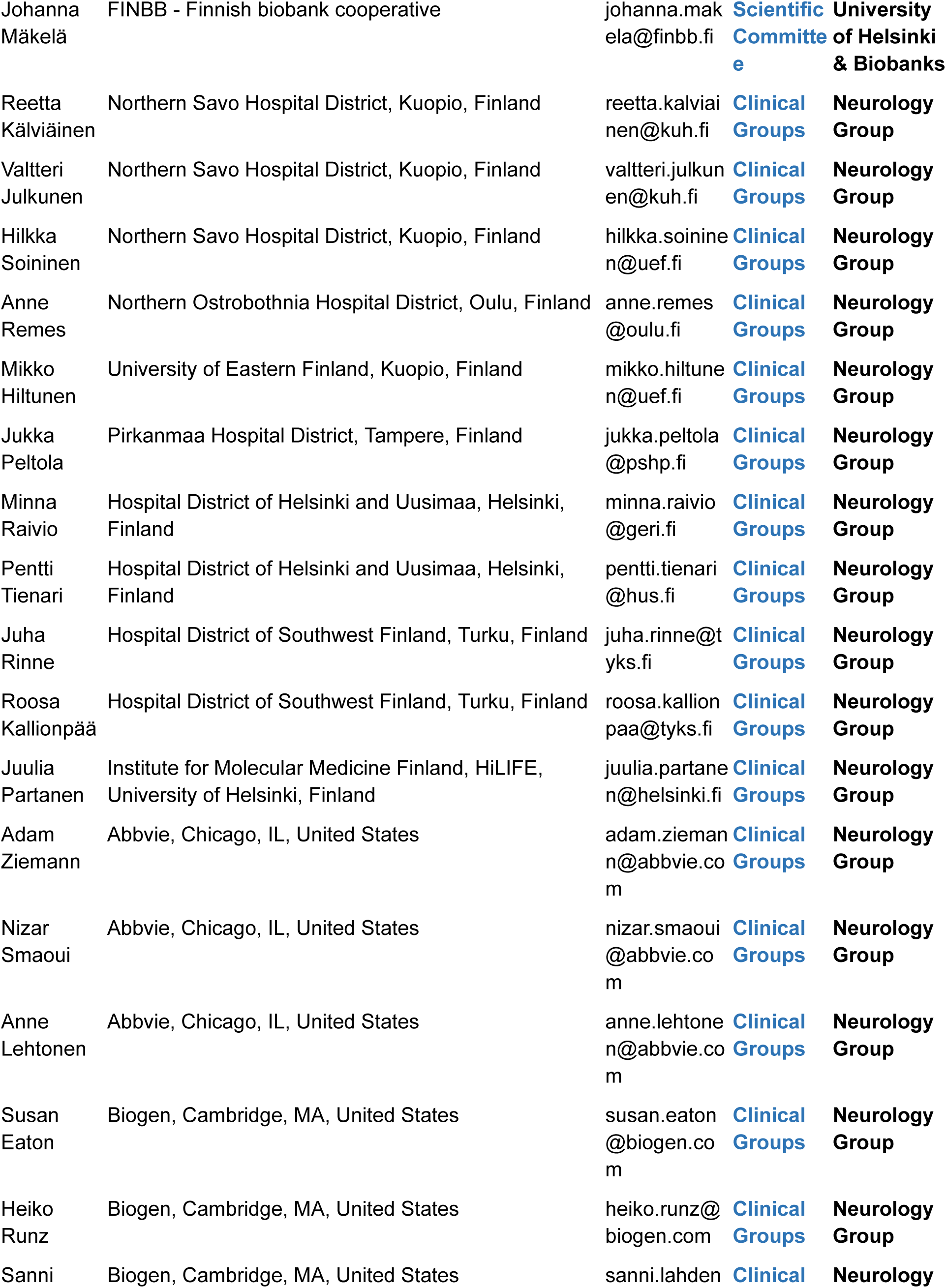
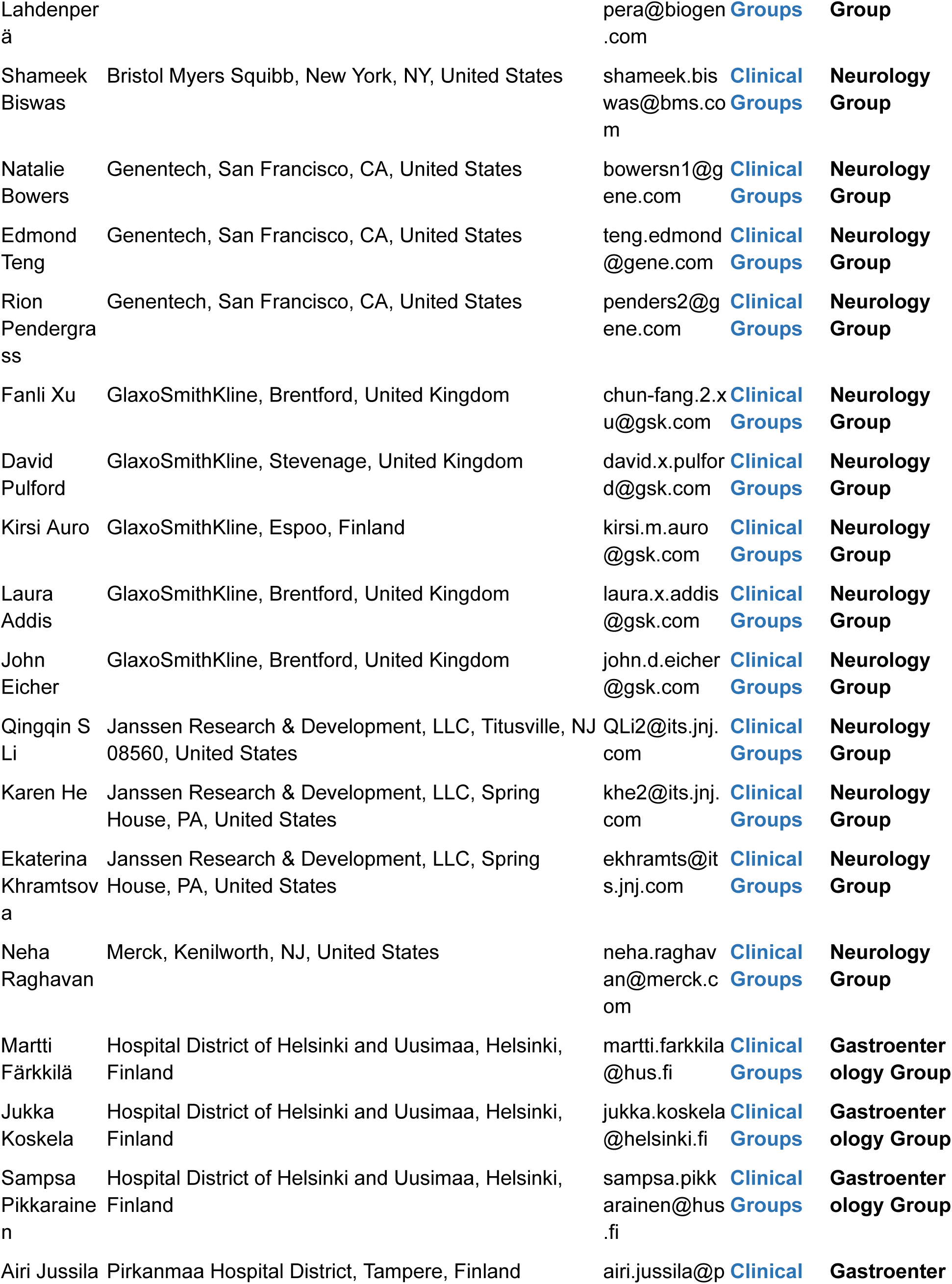
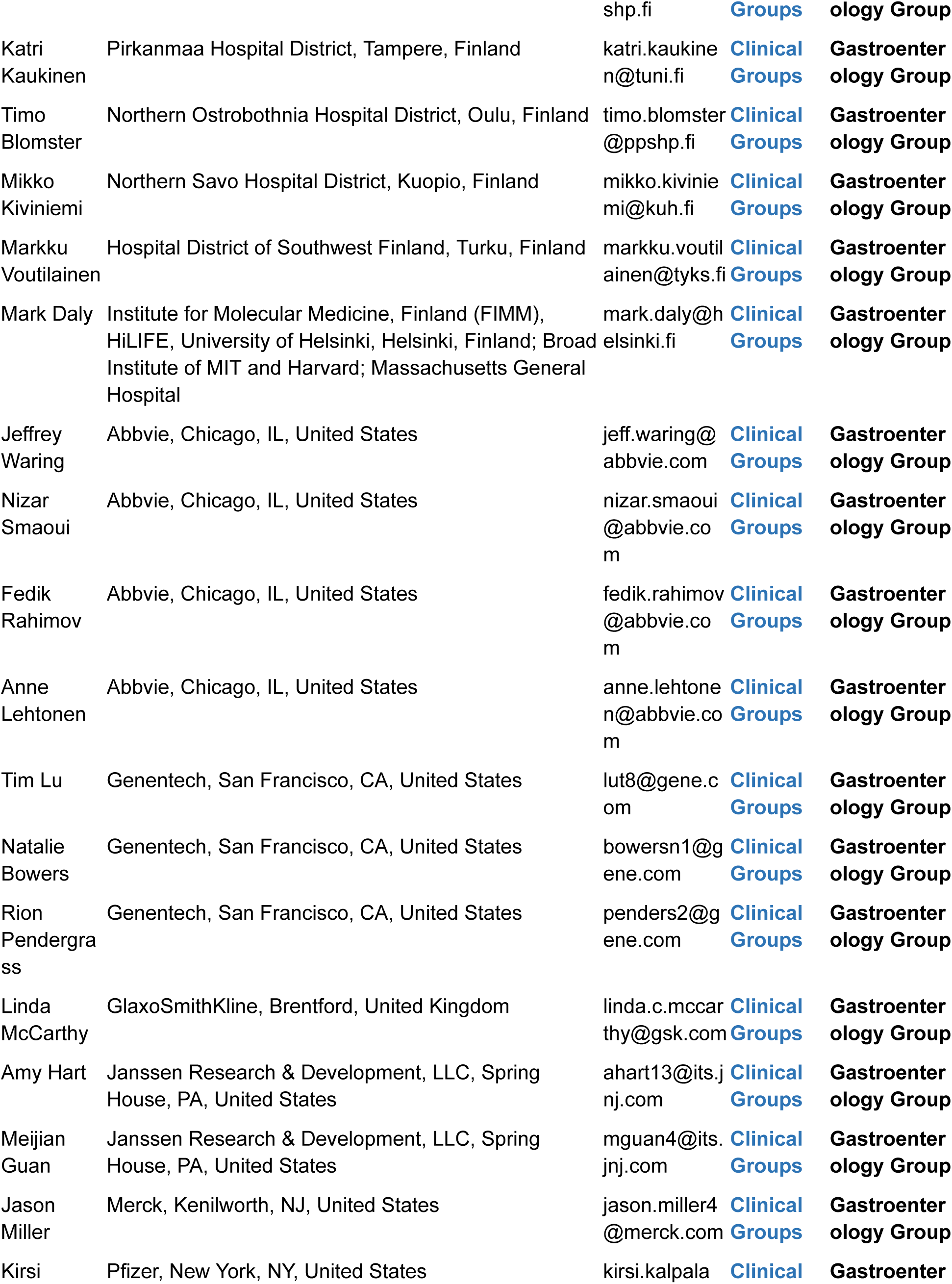
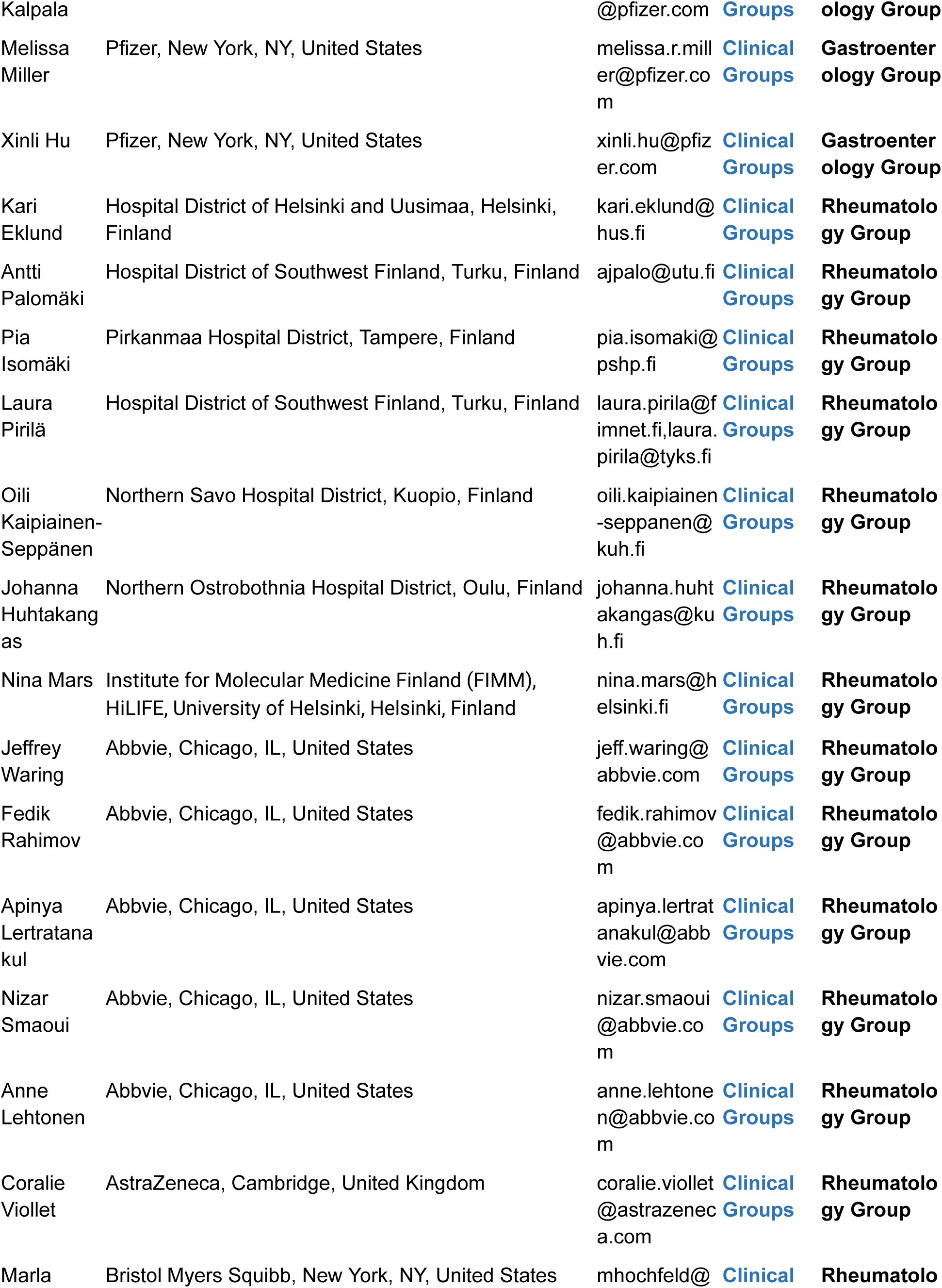
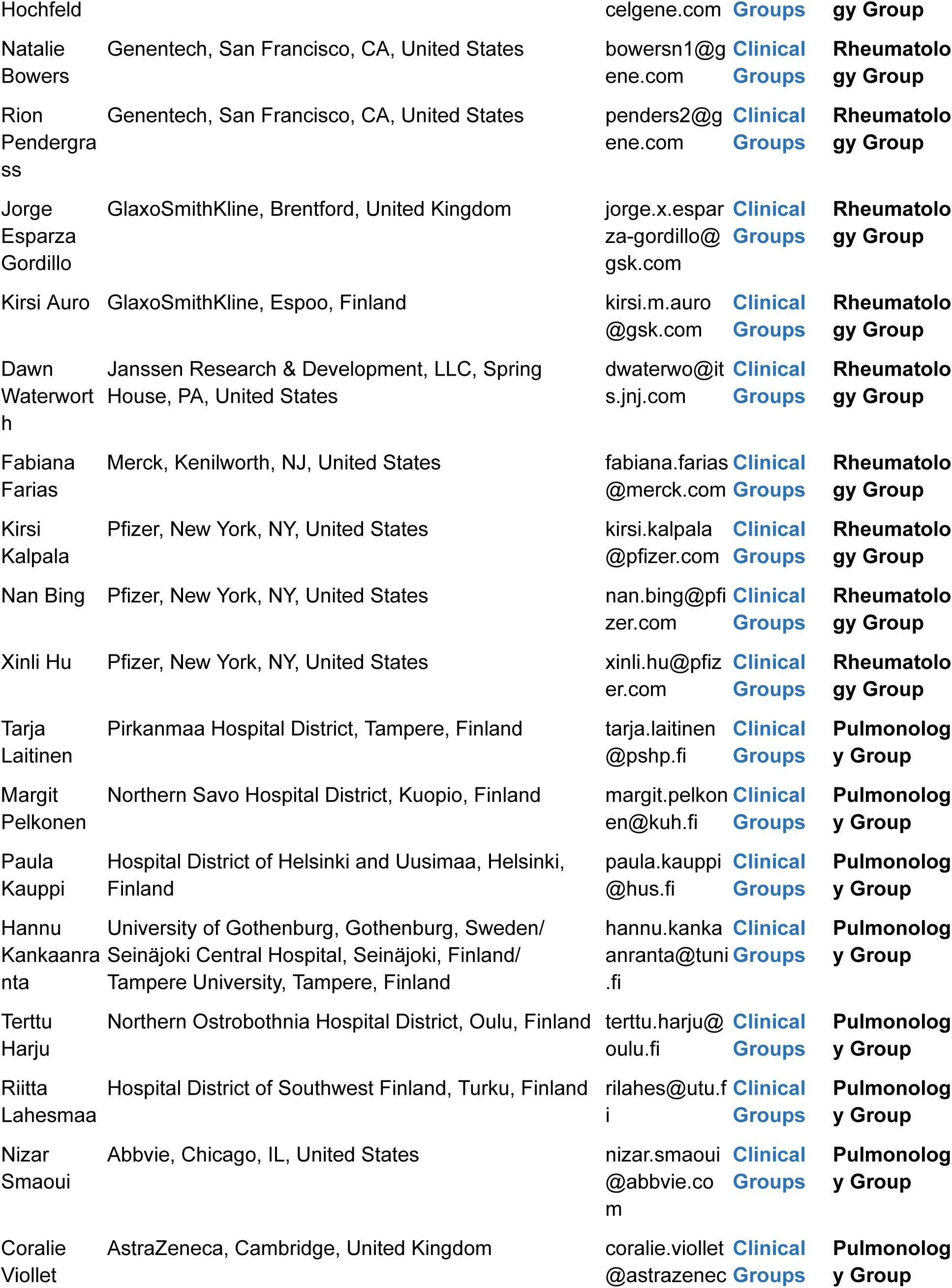
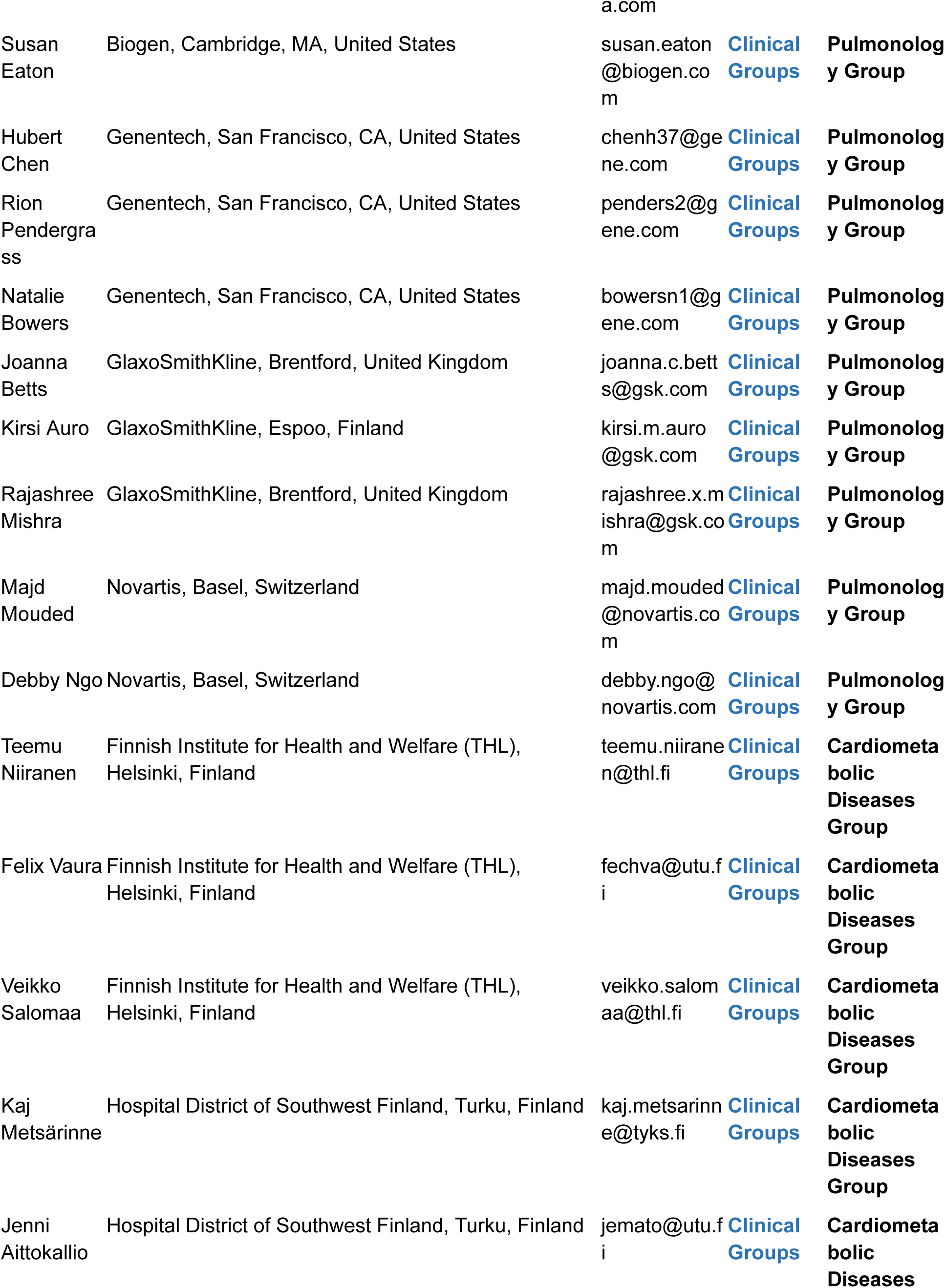
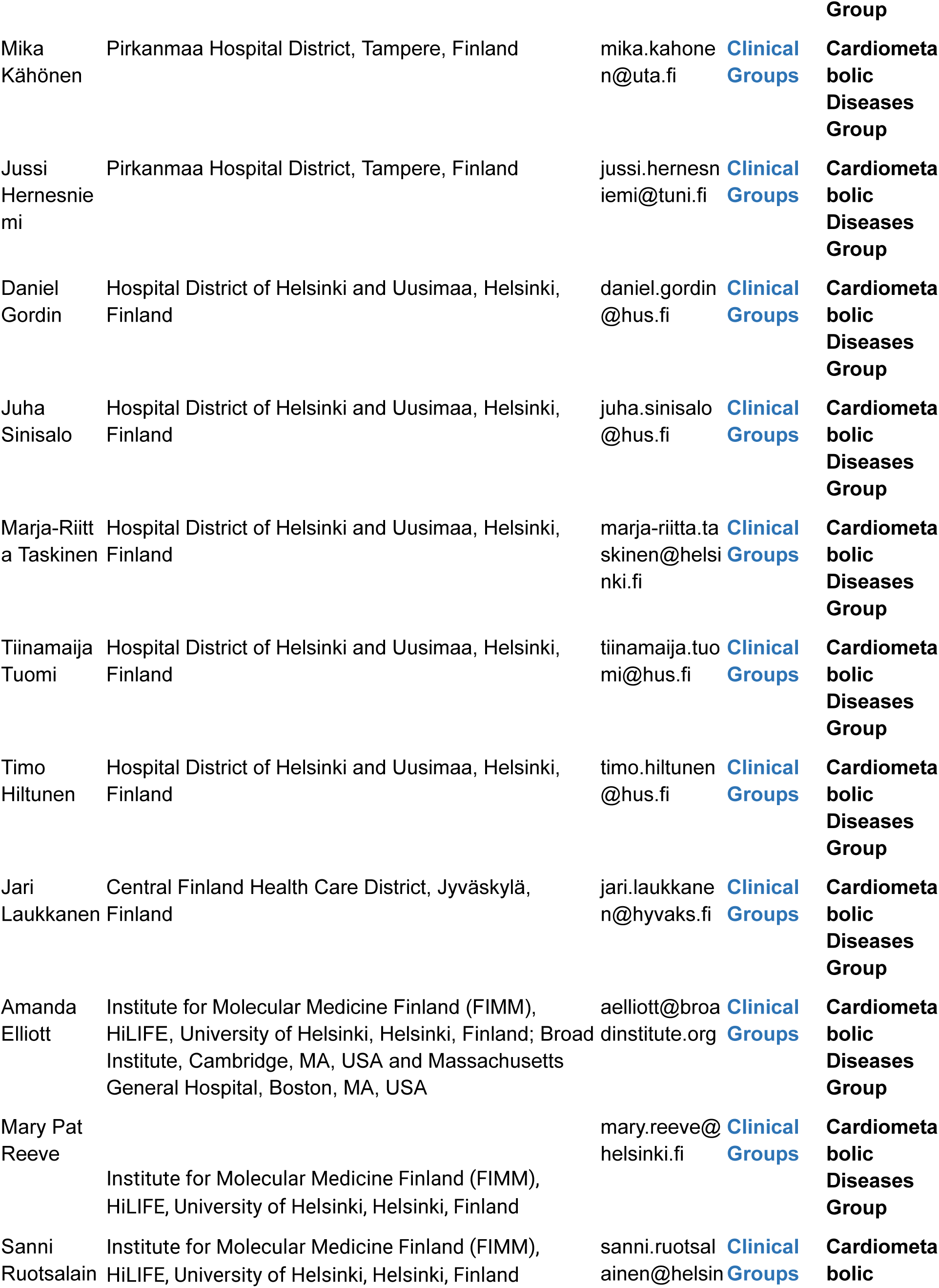
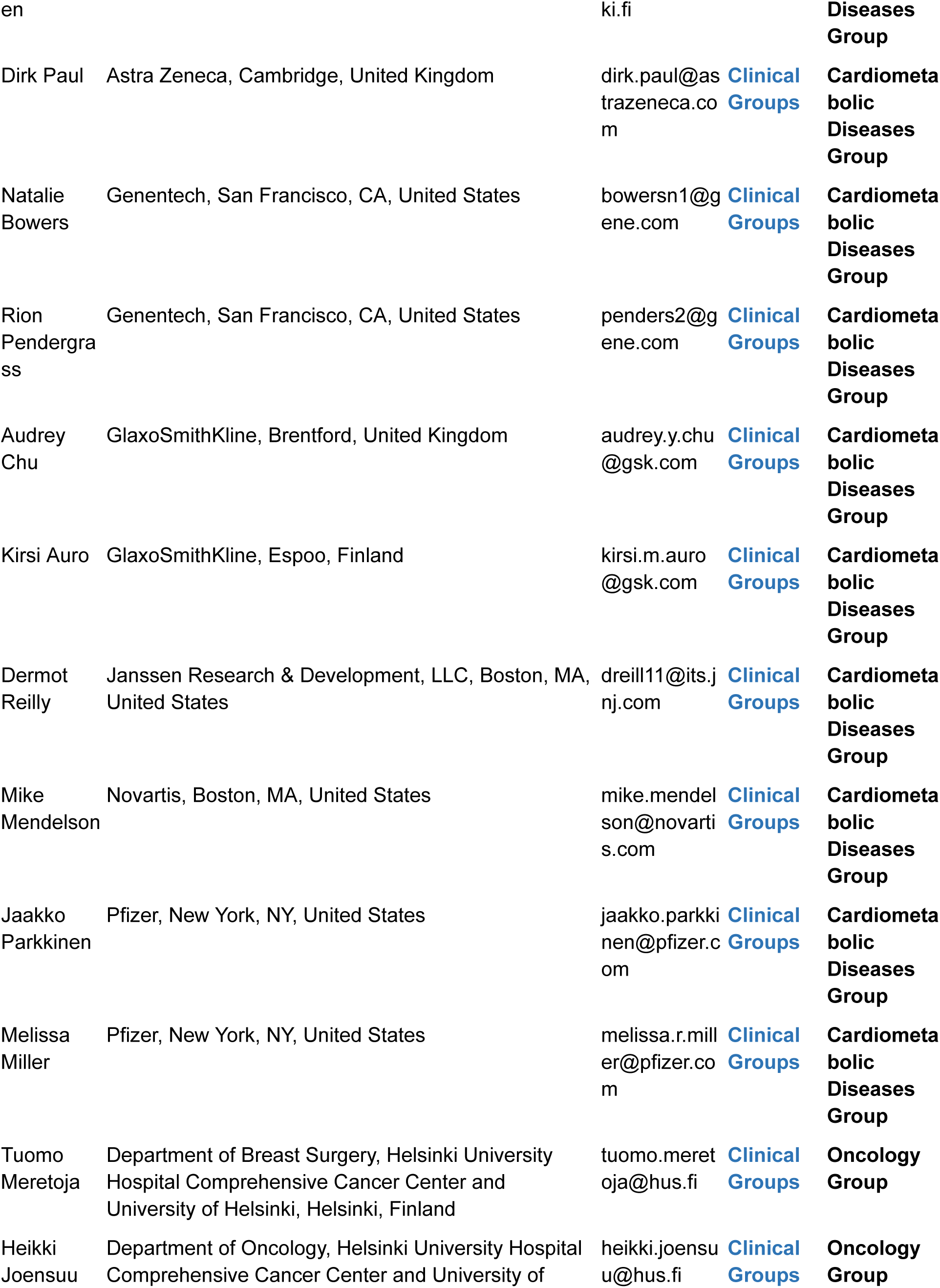
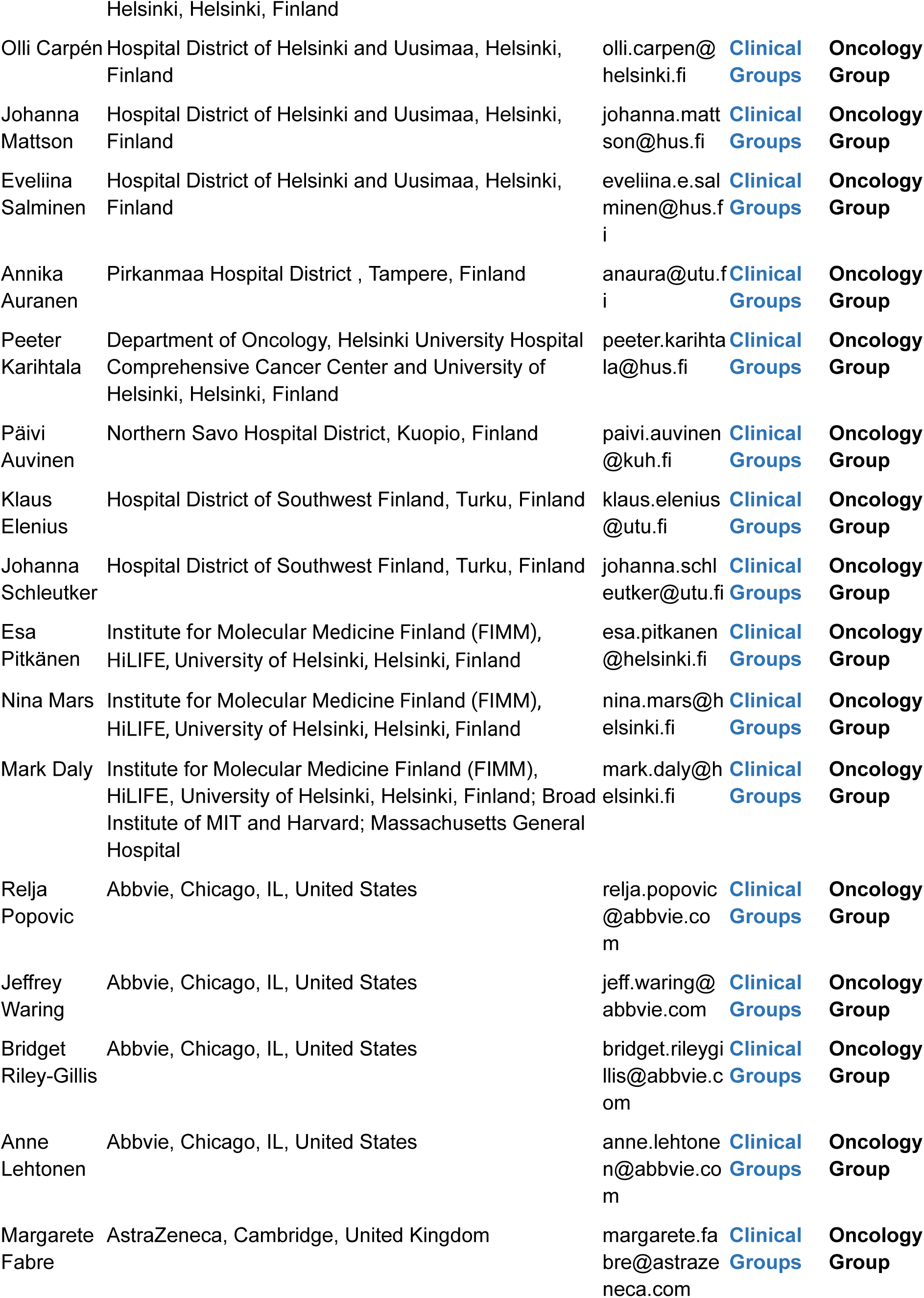
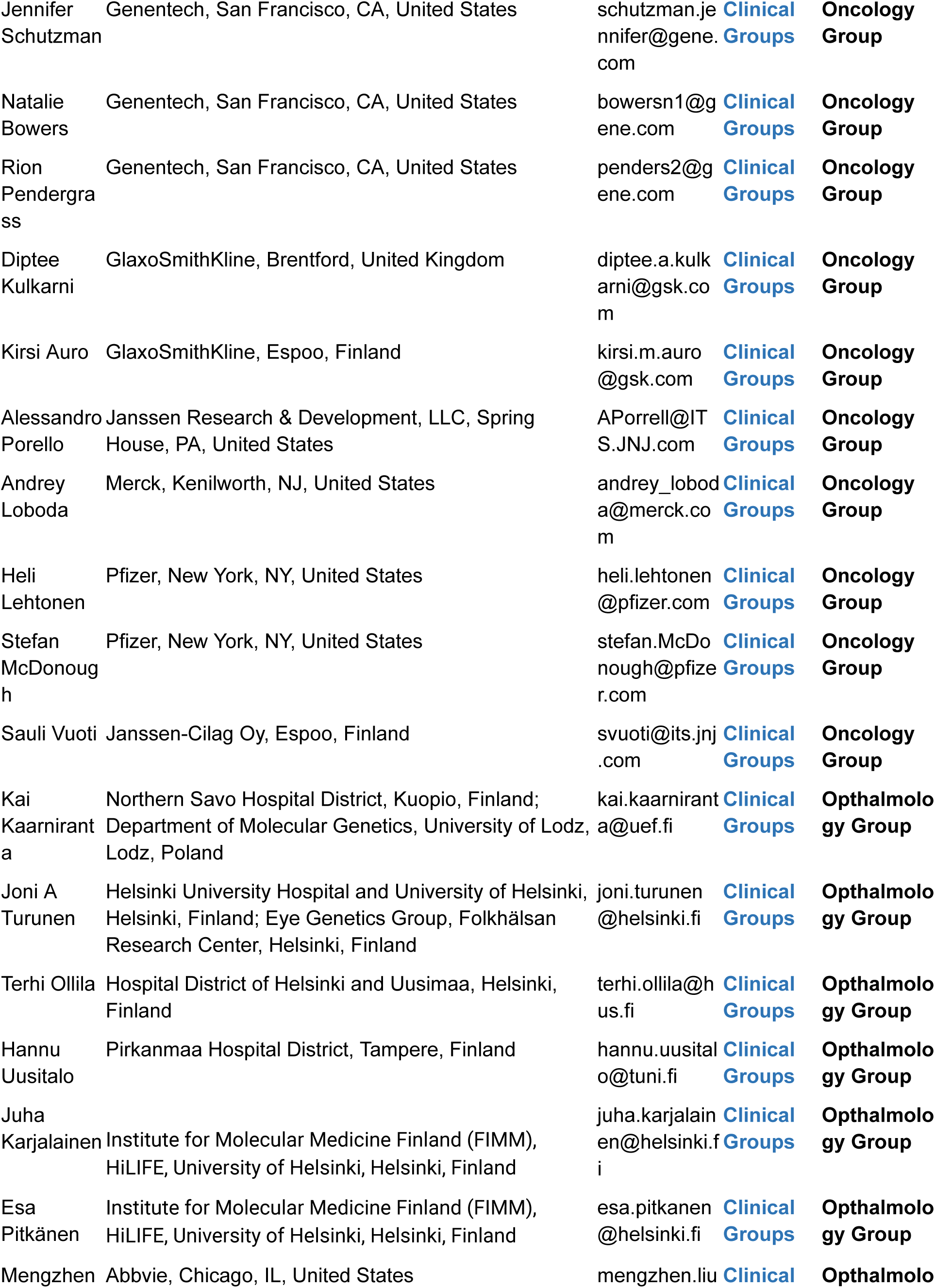
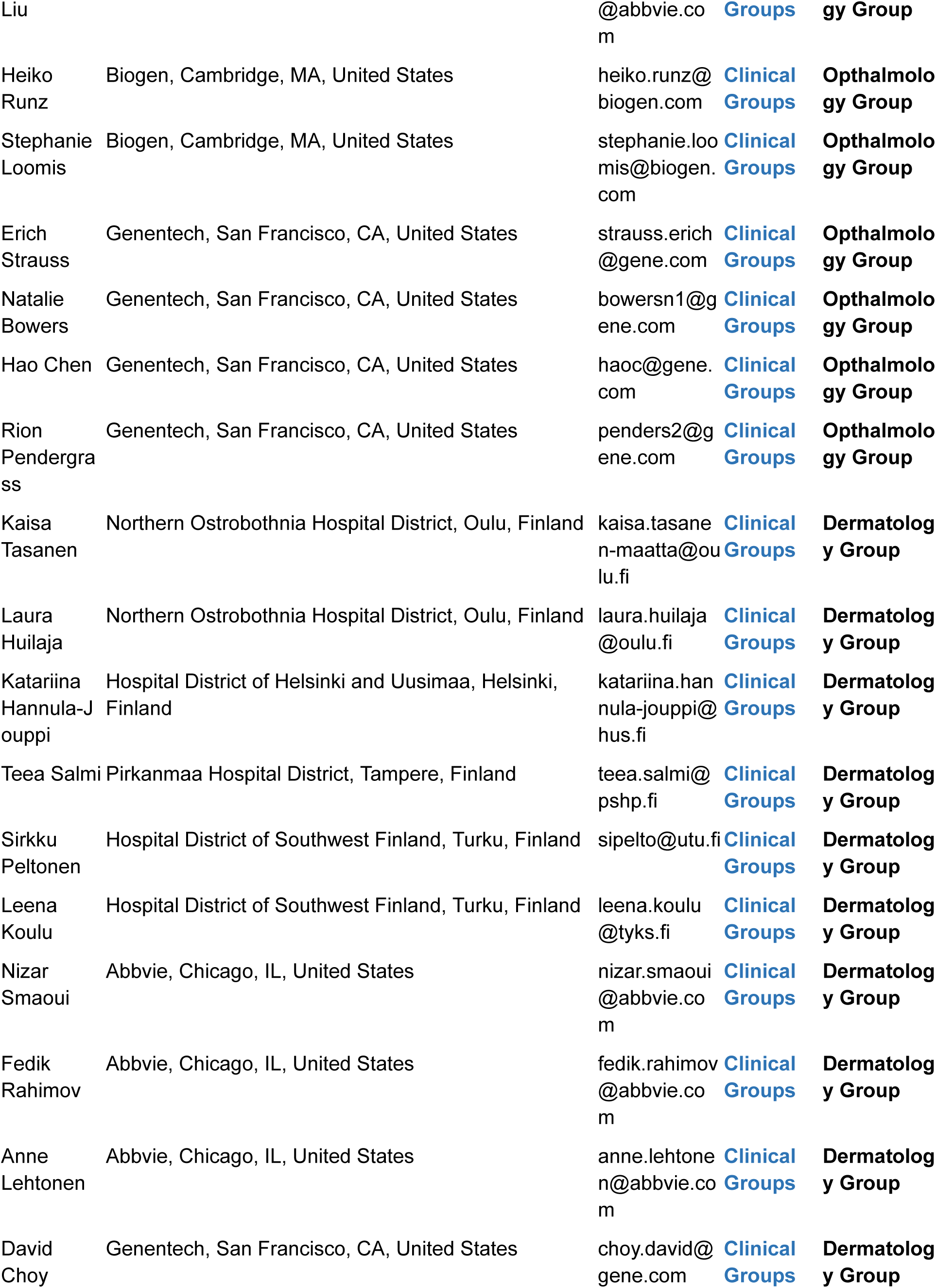
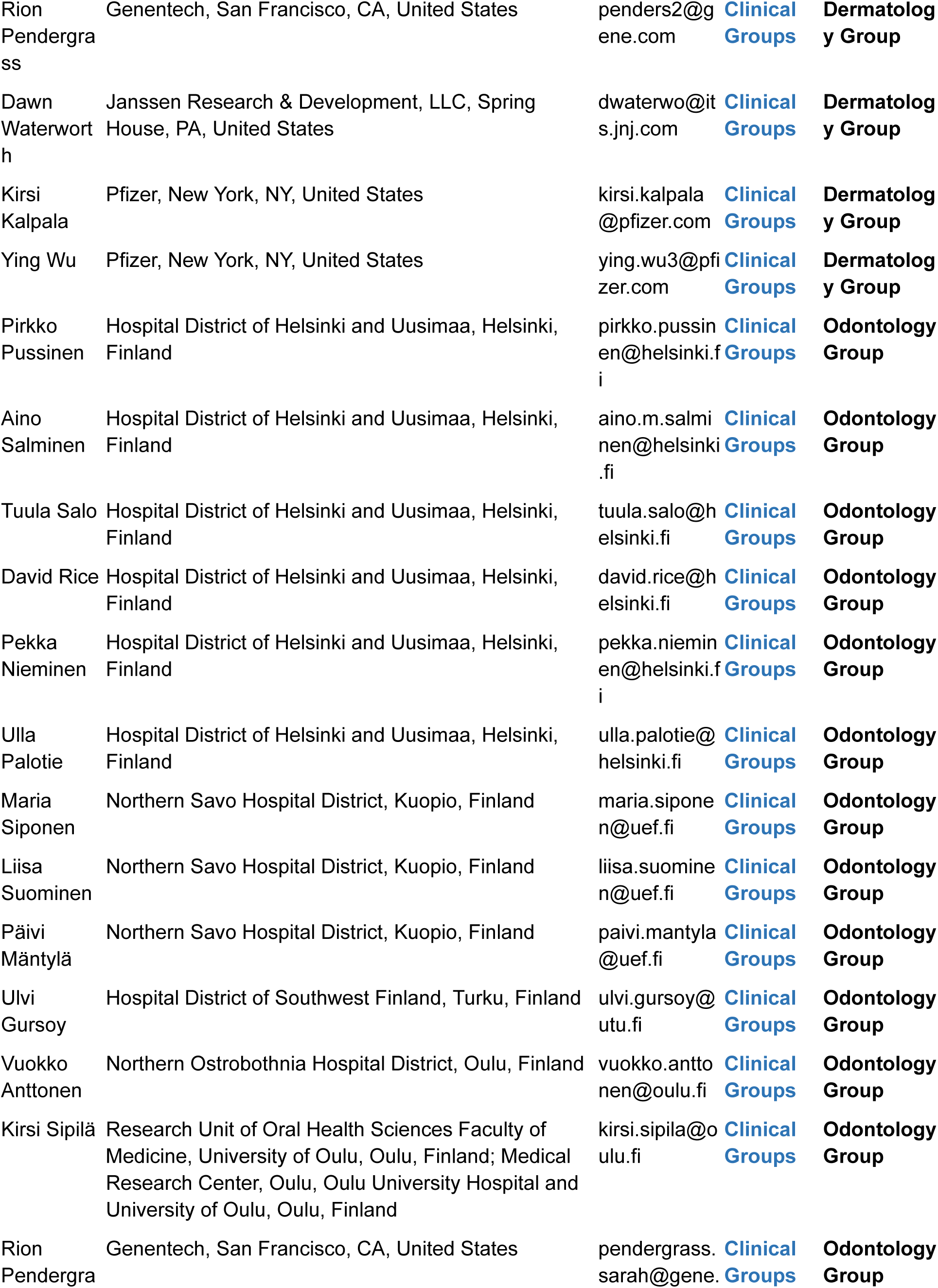
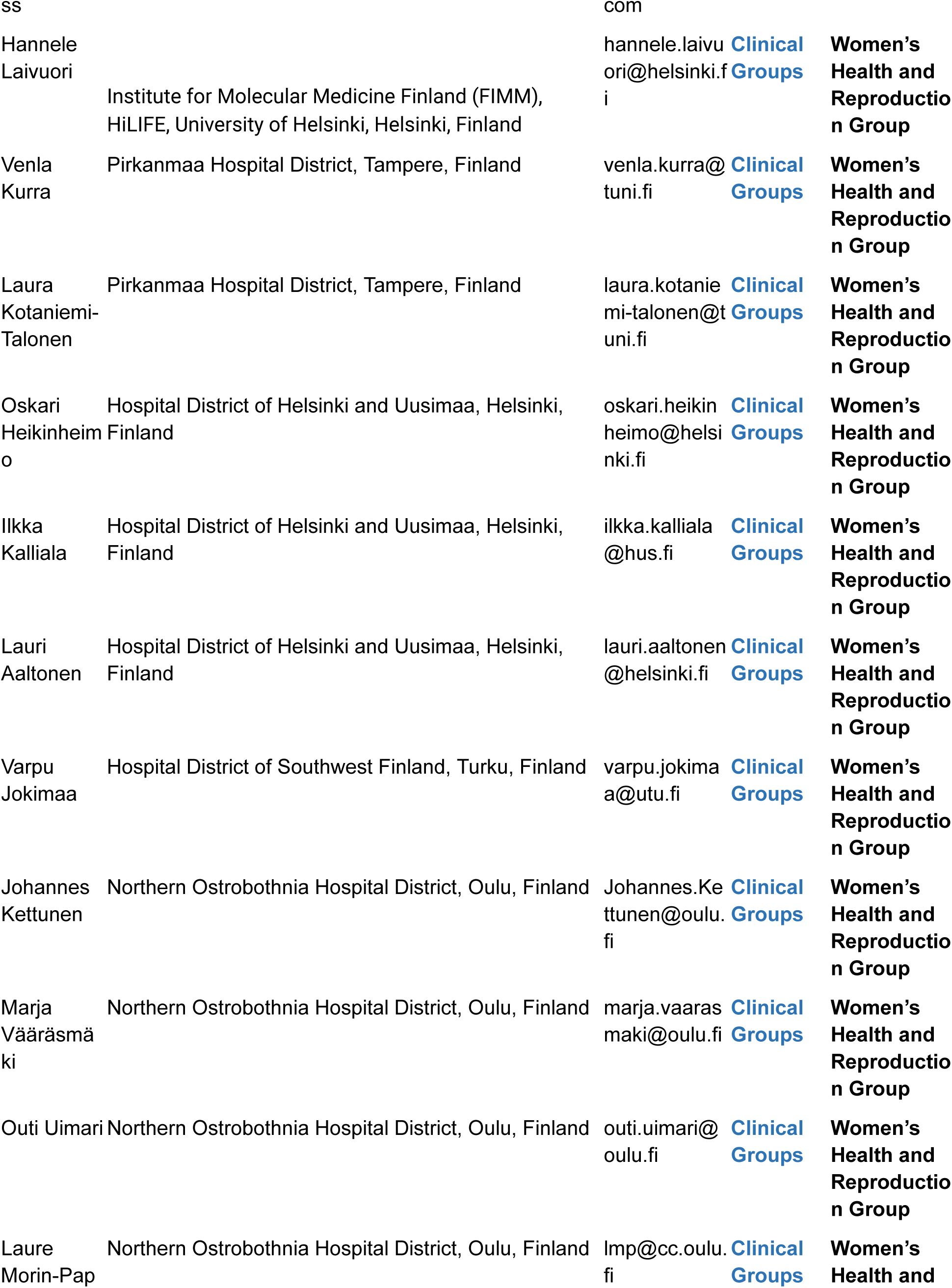
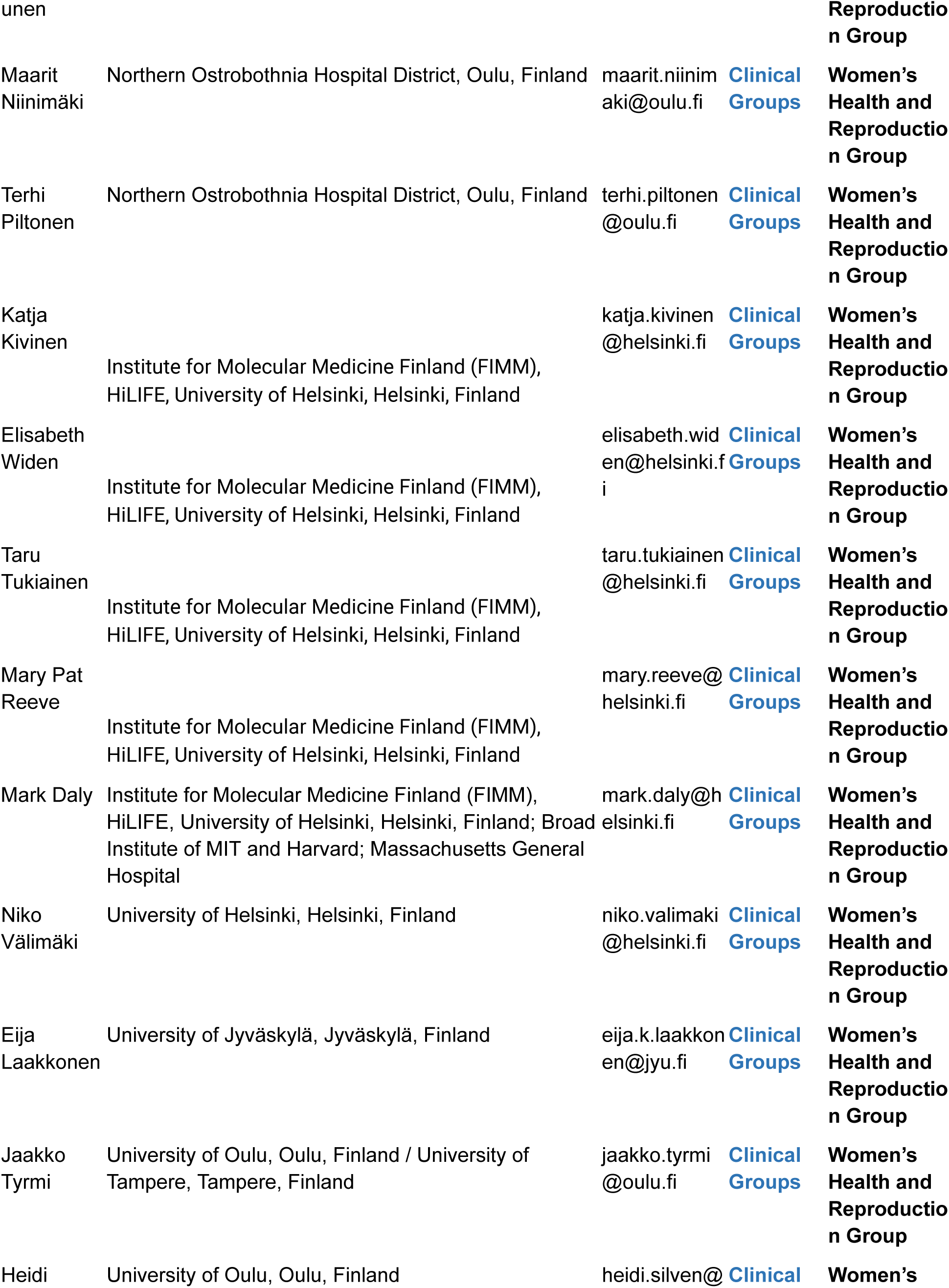
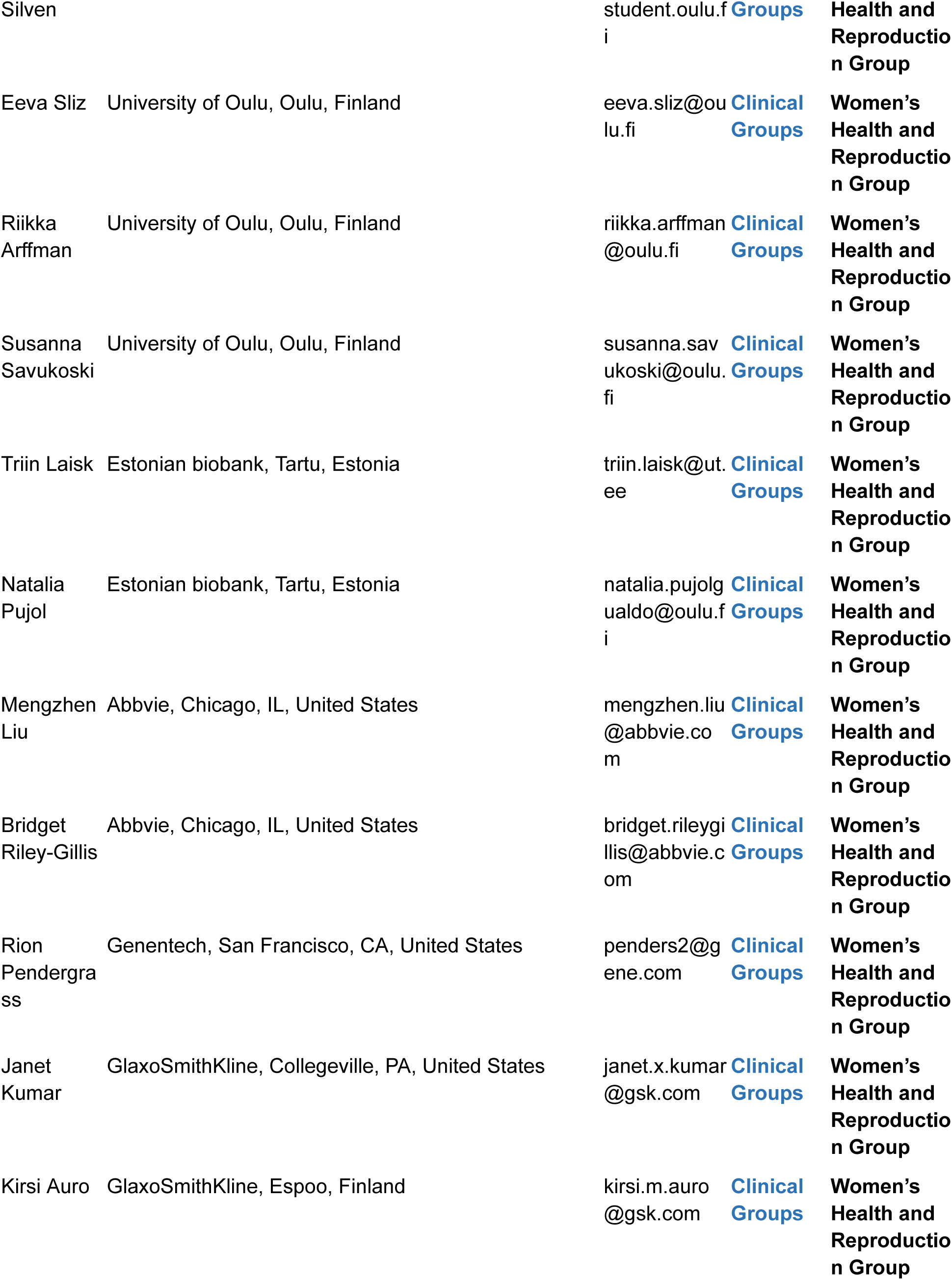
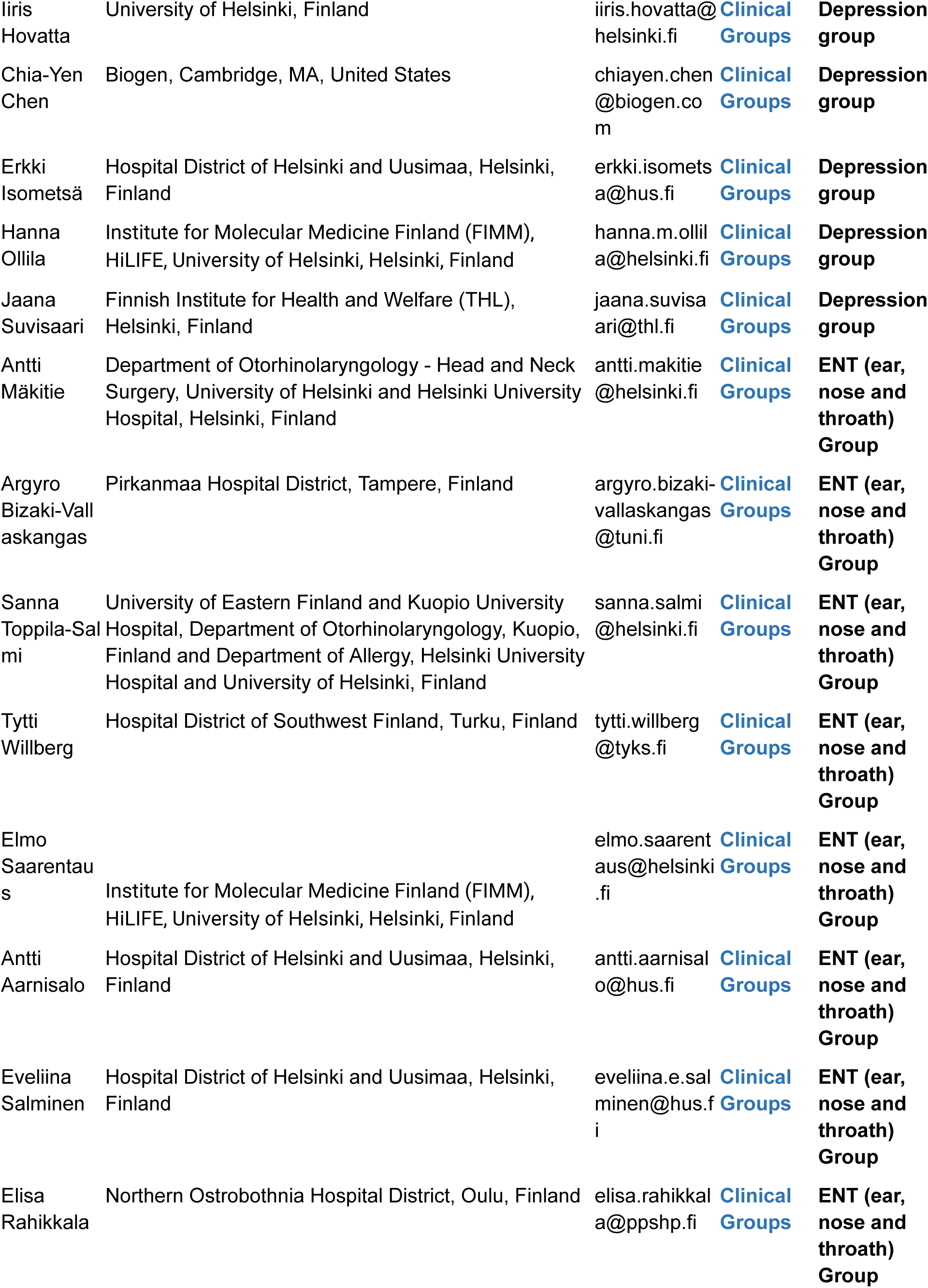
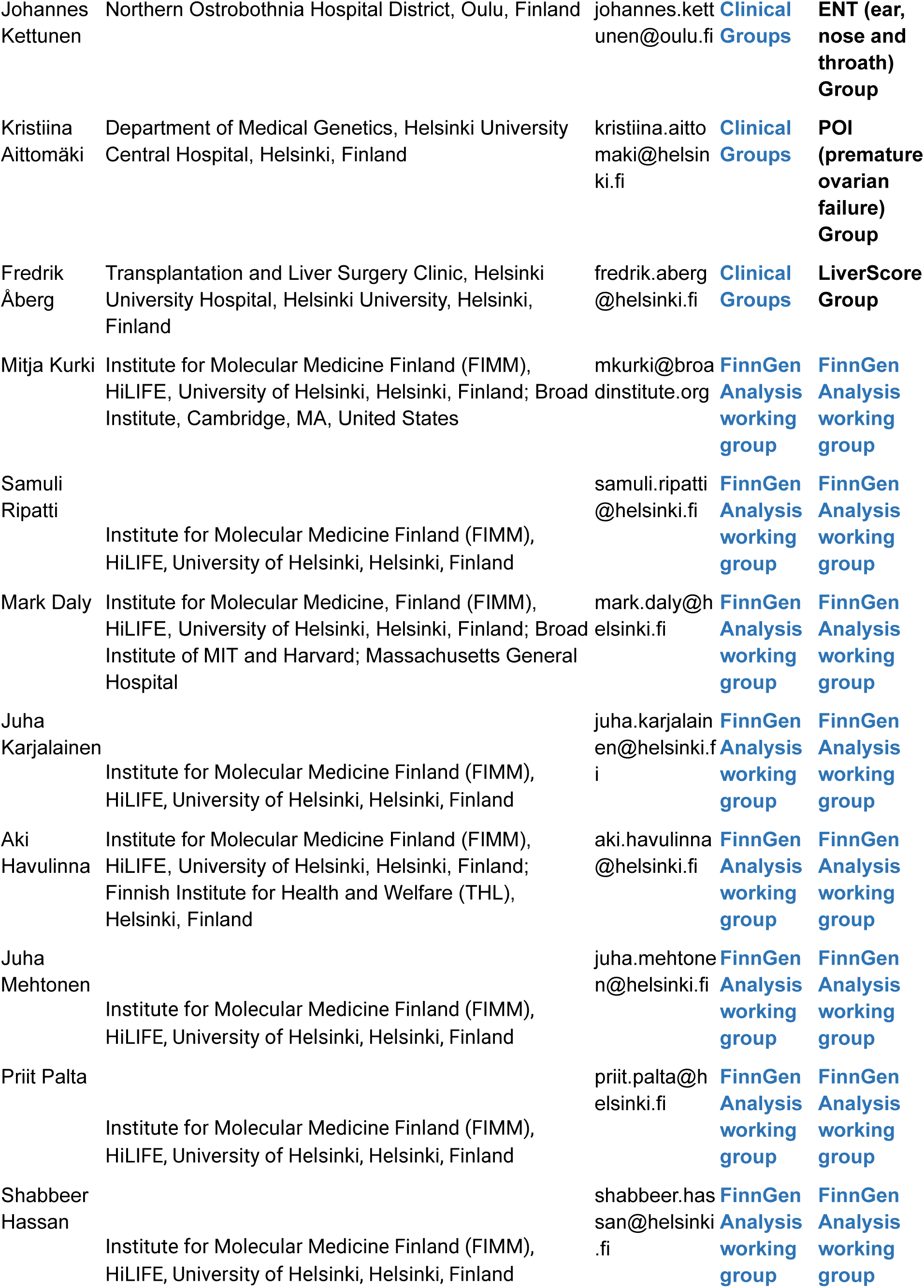
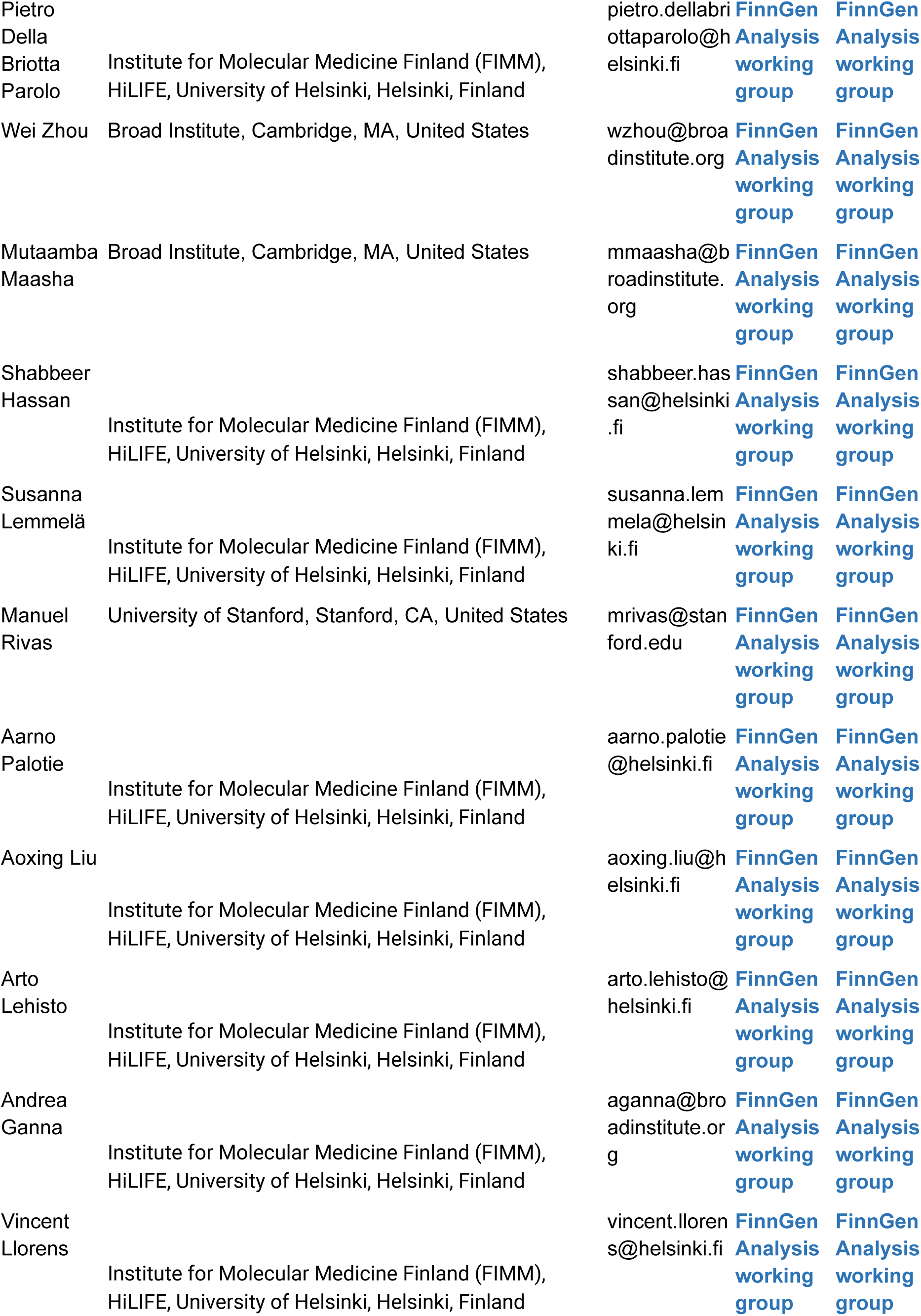
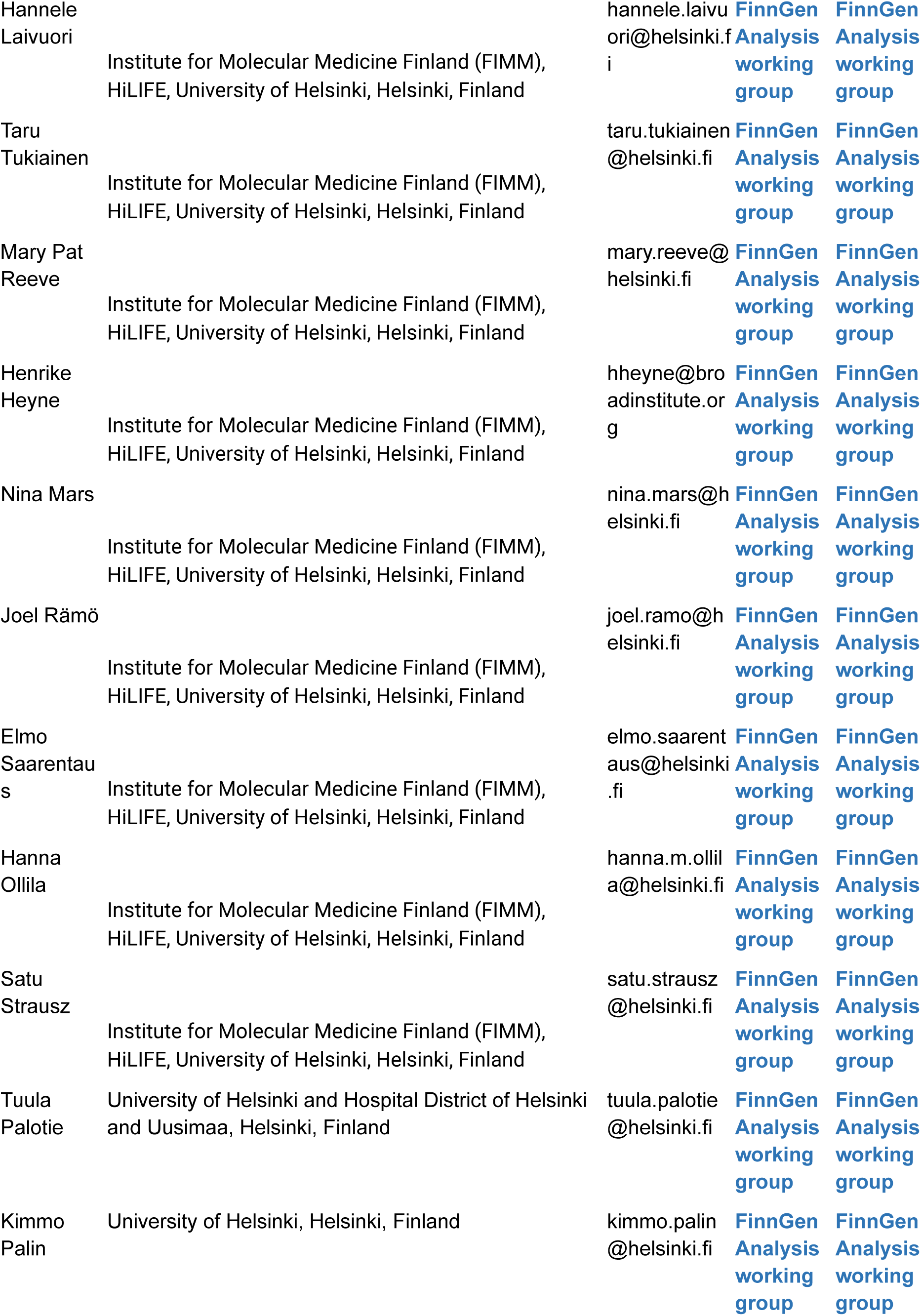
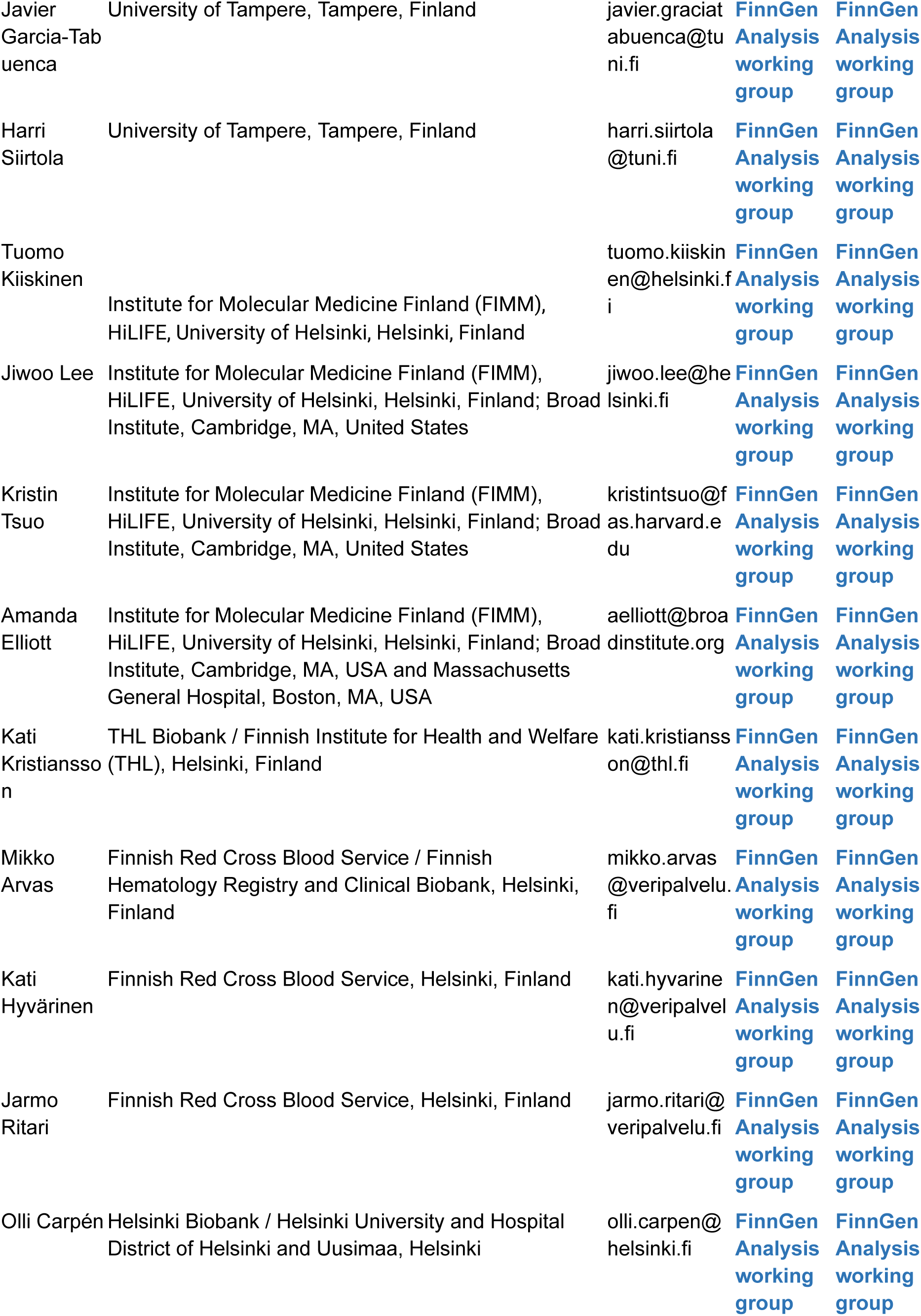
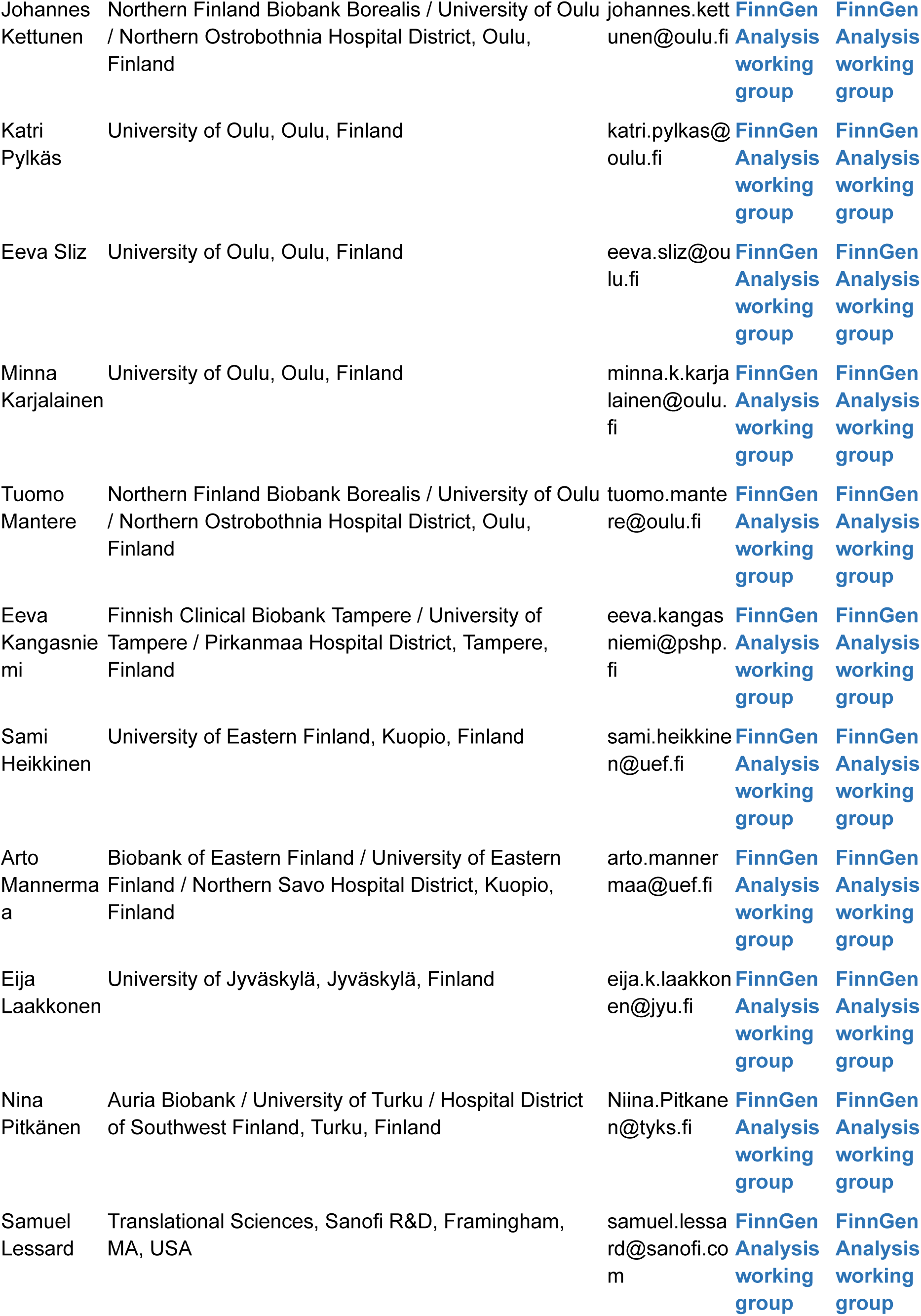
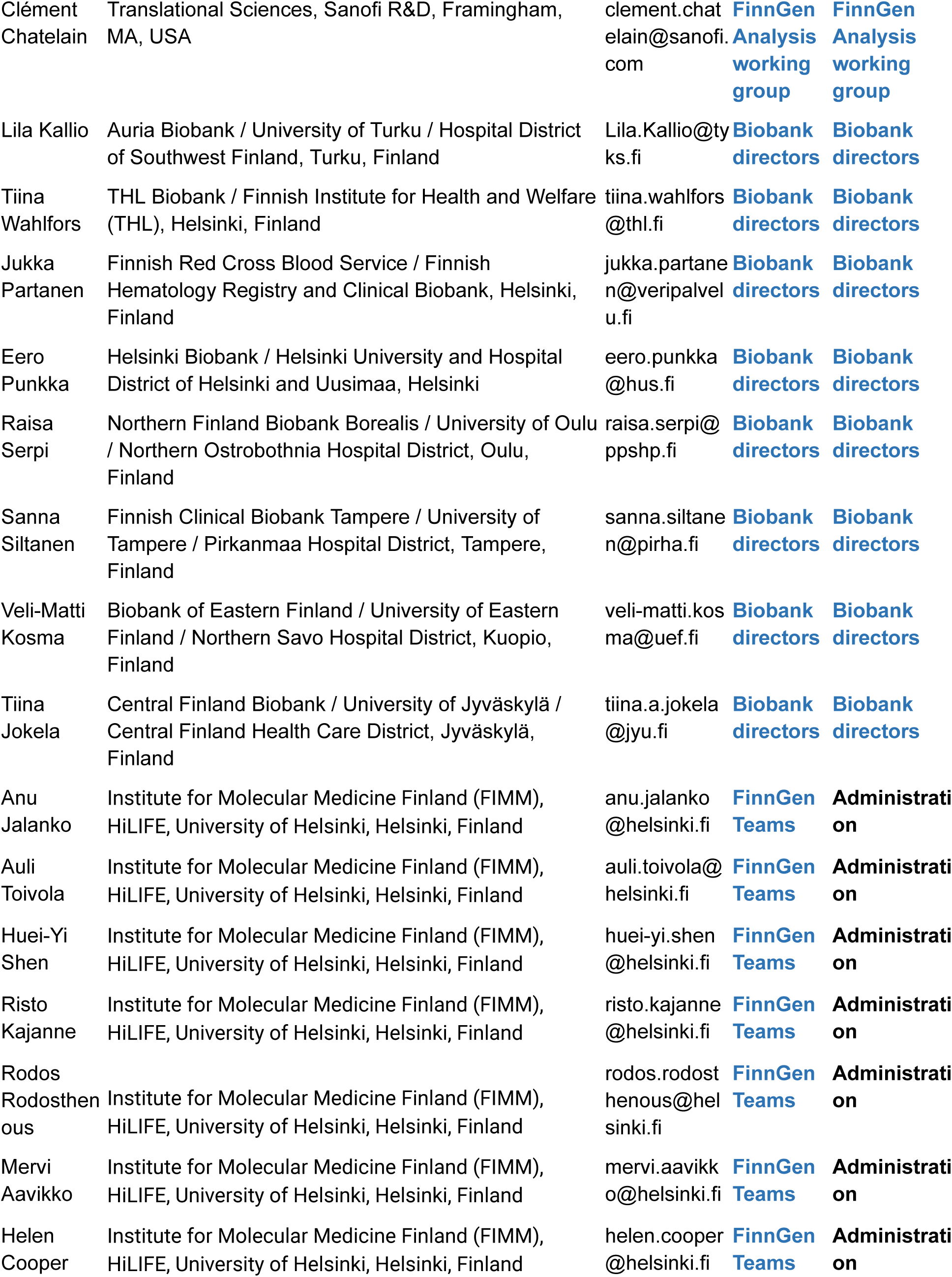
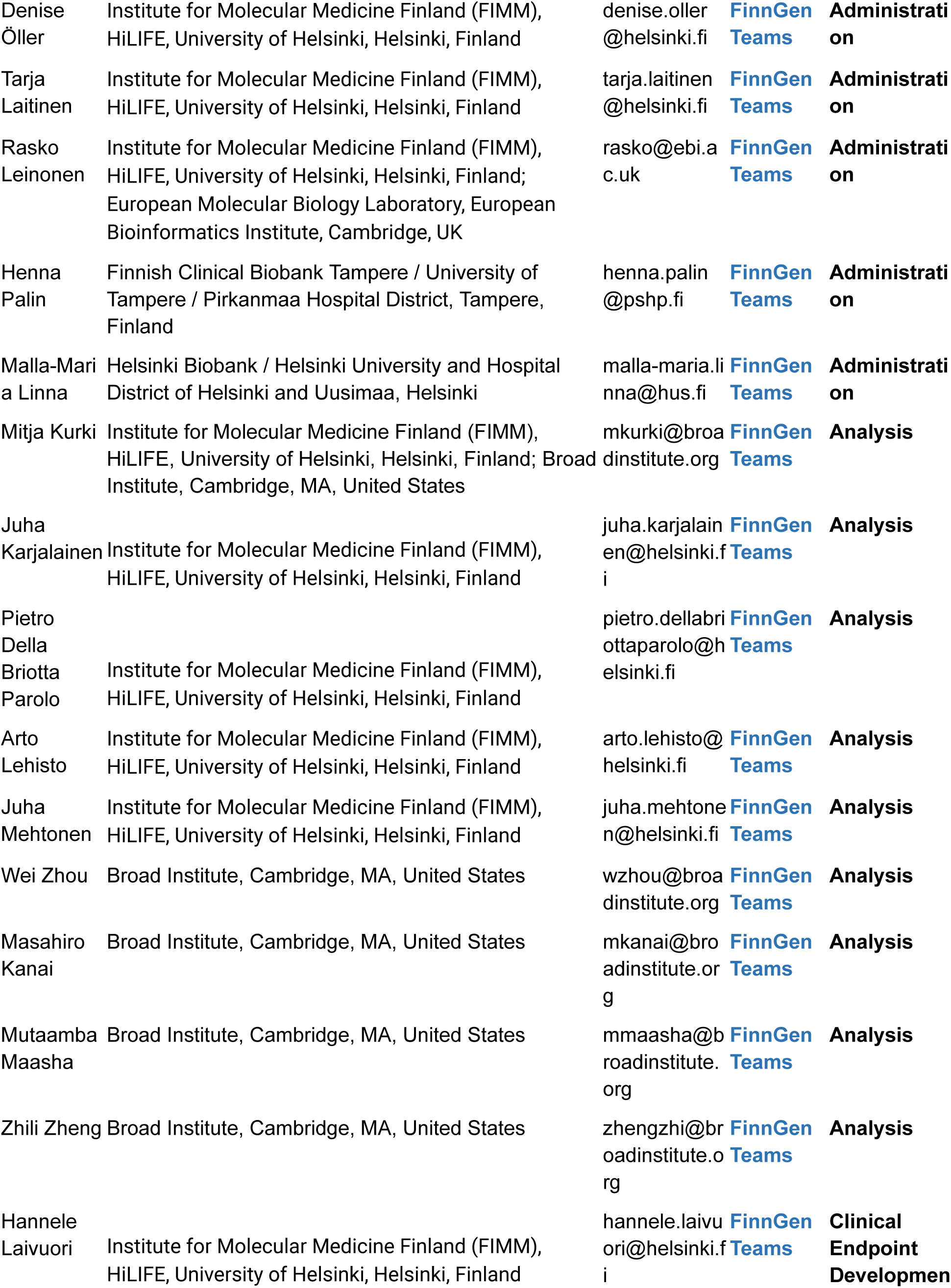
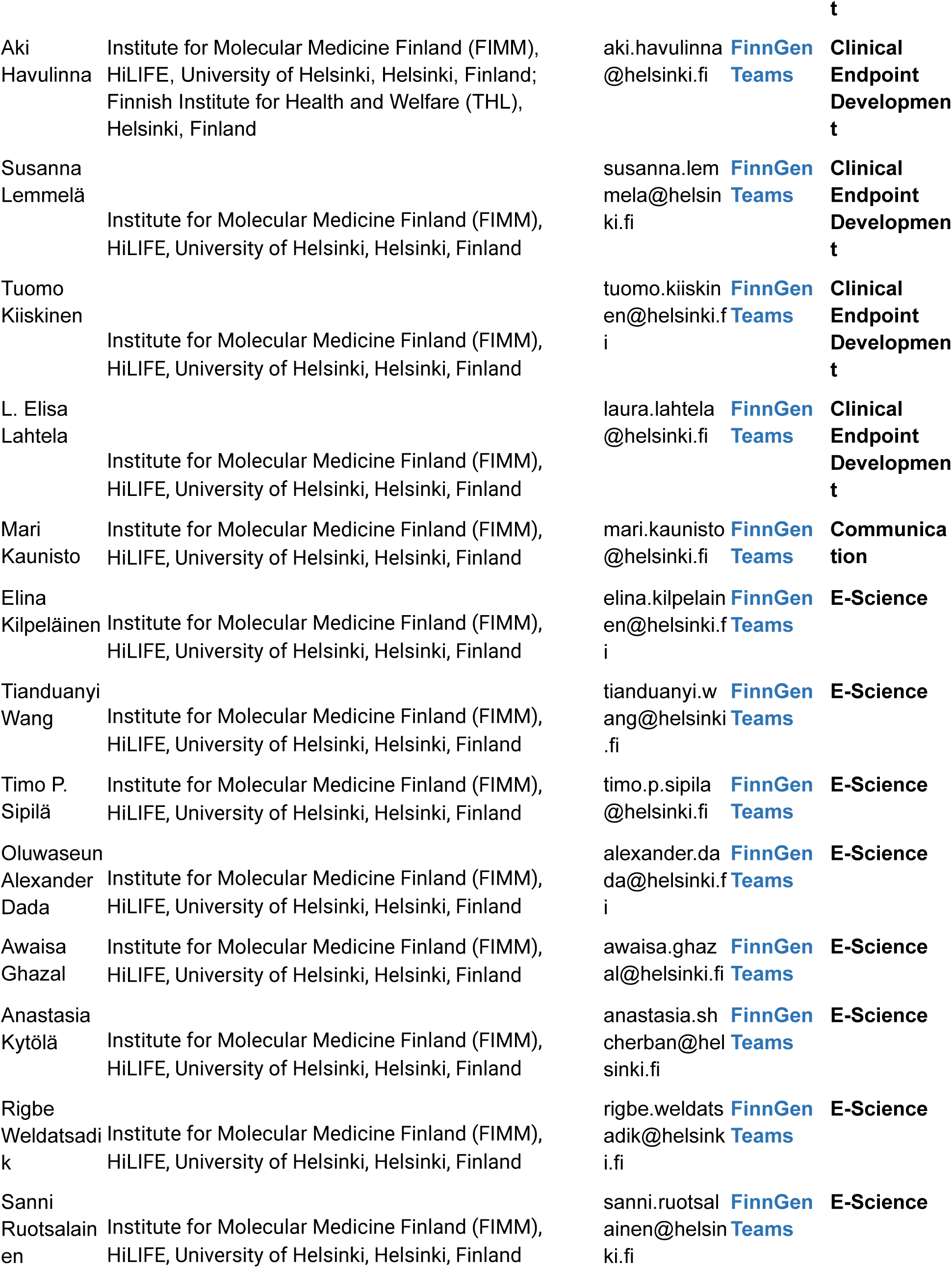
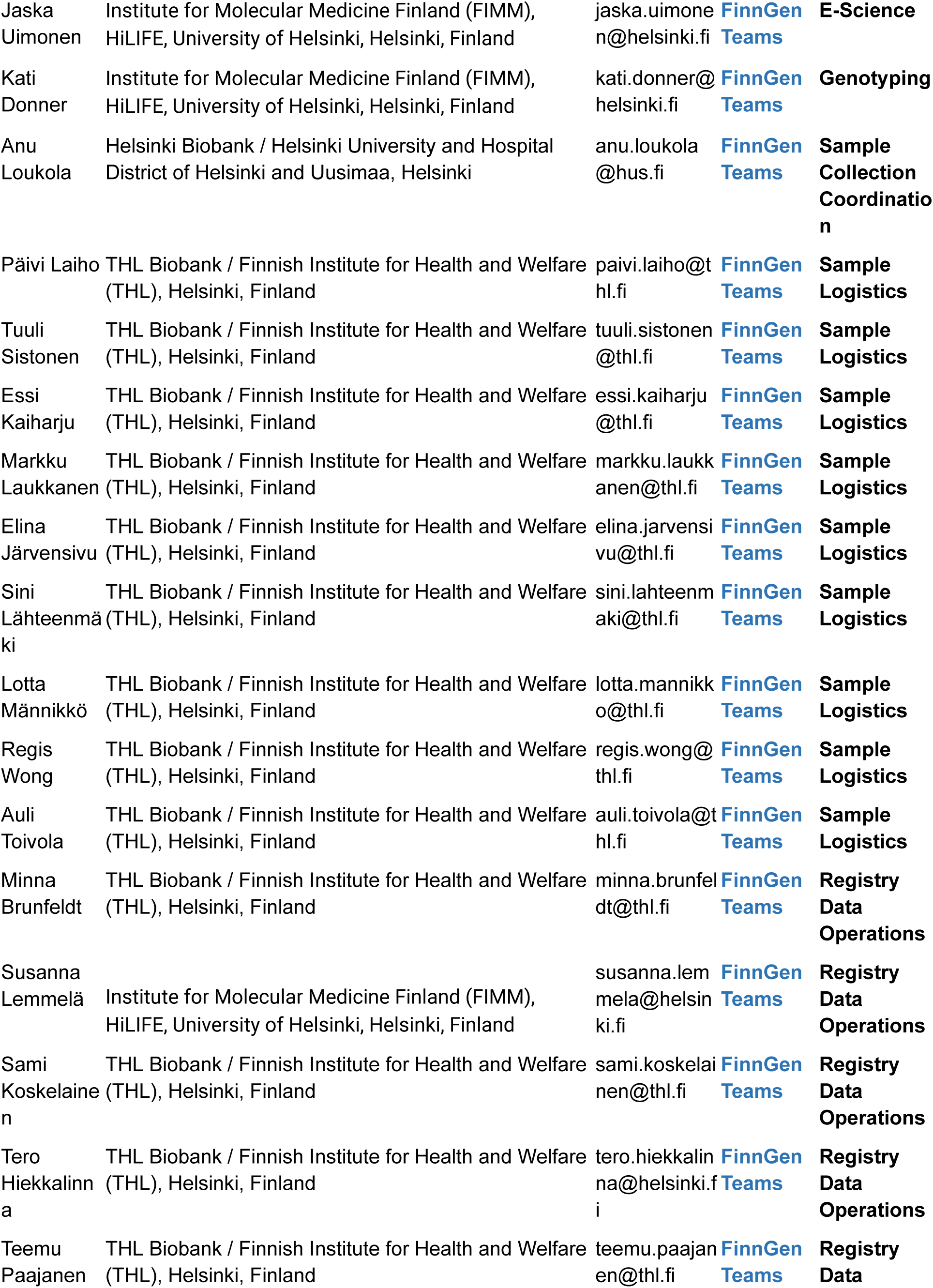
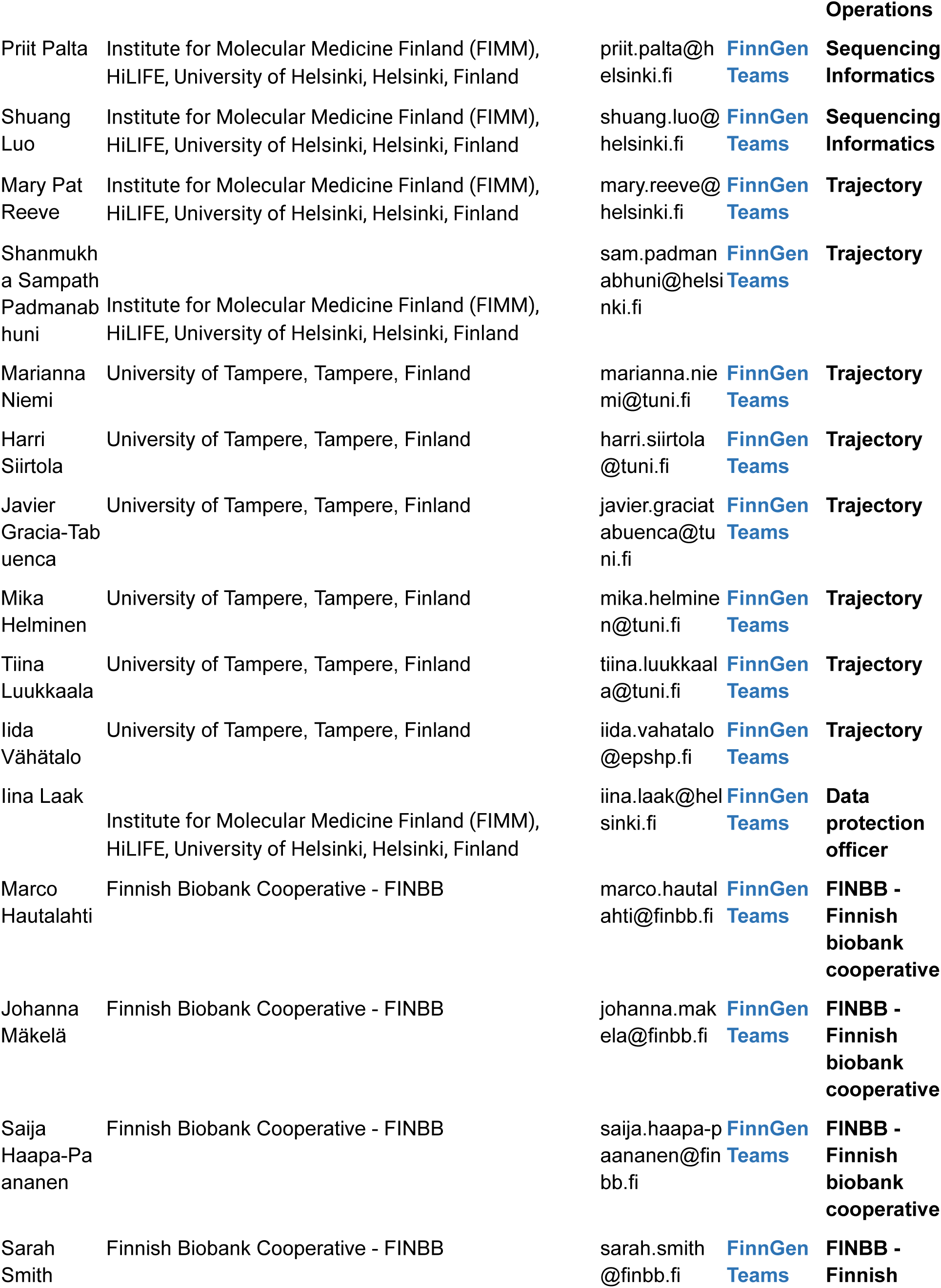
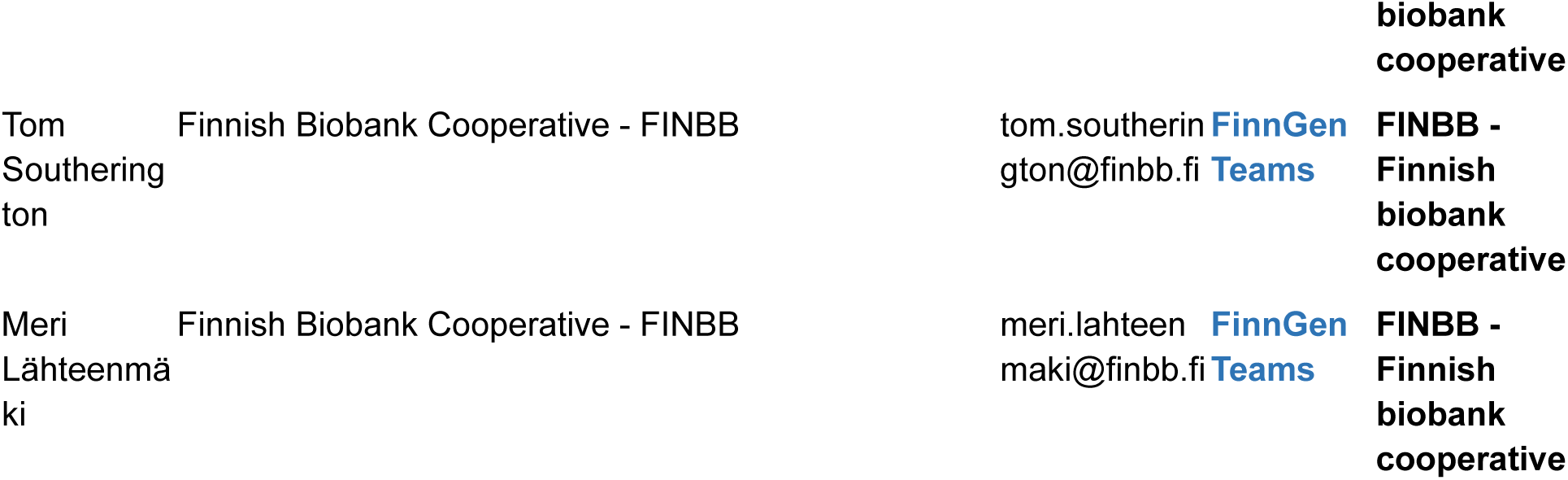

## References

1. Russell, W. R. *The Traumatic Amnesias*. (Oxford University Press, USA, 1971).

2. Ropper, A. H. Transient Global Amnesia. N. Engl. J. Med. 388, 635–640 (2023).

3. Bartsch, T. & Deuschl, G. Transient global amnesia: functional anatomy and clinical implications. Lancet Neurol. 9, 205–214 (2010).

4. Nehring, S. M., Spurling, B. C. & Kumar, A. Transient Global Amnesia. in StatPearls [Internet] (StatPearls Publishing, 2024).

5. Rommens, J. M. et al. Identification of the cystic fibrosis gene: chromosome walking and jumping. Science 245, 1059–1065 (1989).

6. Kurki, M. I. et al. FinnGen provides genetic insights from a well-phenotyped isolated population. Nature 613, 508–518 (2023).

7. Allen, N. E. et al. Prospective study design and data analysis in UK Biobank. Sci. Transl. Med. 16, eadf4428 (2024).

8. Bycroft, C. et al. The UK Biobank resource with deep phenotyping and genomic data. Nature 562, 203–209 (2018).

9. Genomic data in the All of Us Research Program. Nature 627, 340–346 (2024).

10. Bulik-Sullivan, B. et al. An atlas of genetic correlations across human diseases and traits. Nat. Genet. 47, 1236–1241 (2015).

11. Weindrich, D., Jennen-Steinmetz, C., Laucht, M., Esser, G. & Schmidt, M. H. Epidemiology and prognosis of specific disorders of language and scholastic skills. Eur. Child Adolesc. Psychiatry 9, 186–194 (2000).

12. GTEx Consortium. Human genomics. The Genotype-Tissue Expression (GTEx) pilot analysis: multitissue gene regulation in humans. Science 348, 648–660 (2015).

13. Chen, E. Y. et al. Enrichr: interactive and collaborative HTML5 gene list enrichment analysis tool. BMC Bioinformatics 14, 128 (2013).

14. Hu, C. et al. CellMarker 2.0: an updated database of manually curated cell markers in human/mouse and web tools based on scRNA-seq data. Nucleic Acids Res. 51, D870–D876 (2023).

15. Tanigawa, Y. et al. Rare protein-altering variants in ANGPTL7 lower intraocular pressure and protect against glaucoma. PLoS Genet. 16, e1008682 (2020).

16. Sun, B. B. et al. Genetic associations of protein-coding variants in human disease. Nature 603, 95–102 (2022).

17. O’Dell, B. L. Cysteine-rich intestinal protein (CRIP): a new intestinal zinc transport protein. Nutr. Rev. 50, 232–233 (1992).

18. Zhang, L. et al. Cysteine-rich intestinal protein 1 suppresses apoptosis and chemosensitivity to 5-fluorouracil in colorectal cancer through ubiquitin-mediated Fas degradation. J. Exp. Clin. Cancer Res. 38, 120 (2019).

19. Kanai, M. et al. Meta-analysis fine-mapping is often miscalibrated at single-variant resolution. Cell Genom 2, (2022).

